# Determinants of Access to Autism Spectrum Disorder Diagnostic Services: A Systematic Review and Meta-analysis of Factors Associated with Diagnostic Completion, Diagnostic Pathways, and Timely Diagnosis

**DOI:** 10.64898/2026.08.26.26361221

**Authors:** John K. Muthuka, Lispah Wanjiku, Chrisphine Onyango, Kelly Oluoch, Mary T. Kioko, Jacinta Maluki, Japheth M. Nzioki, Sara Kim

## Abstract

**Background:** Autism spectrum disorder (ASD) is a lifelong neurodevelopmental condition for which timely diagnosis is critical to early intervention, family support, and equitable access to care. However, substantial disparities in access to ASD diagnostic services persist across socioeconomic, geographic, clinical, and health-system contexts. This systematic review and meta-analysis synthesized evidence on determinants of access across the ASD diagnostic pathway, from recognition and referral to diagnostic completion and timely diagnosis.

**Methods:** We systematically searched MEDLINE/PubMed, Embase, Scopus, Web of Science, Global Health, and grey-literature sources for studies published between January 2004 and December 2024. Eligible studies examined determinants of ASD diagnostic completion, diagnostic pathways, diagnostic timeliness, or barriers and facilitators to diagnostic access. Two reviewers independently extracted data and assessed methodological quality using the Mixed Methods Appraisal Tool (MMAT). Quantitatively comparable estimates were synthesized using random-effects models with restricted maximum likelihood estimation. Heterogeneity was assessed using Cochran’s Q, I2, tau2, and 95% prediction intervals. Pre-specified subgroup analyses, meta-regression, sensitivity analyses, funnel-plot assessments, and Bayesian random-effects analyses were undertaken.

**Results:** The search identified 4,899 records; after removal of 537 records without associated data, 4,362 records underwent title/abstract screening. 3,800 records were excluded, 562 reports were sought for retrieval, and 450 full-text reports were assessed after 112 could not be retrieved. Ultimately, 22 unique studies met the inclusion criteria. Nine unique studies contributed 23 quantitative effect estimates, while the remaining studies contributed to the narrative synthesis. The evidence covered socioeconomic, geographic, family, communication, screening, child developmental, provider, and health-system determinants. The overall random-effects meta-analysis yielded a pooled diagnostic access outcome of 74.1% (95% CI 65.8-81.1%), with substantial heterogeneity (Qe=209.95, p<0.001; I2=88.4%, 95% CI 79.1-94.4%; tau2=0.691) and a wide 95% prediction interval of 32.8-94.4%. Bayesian analysis produced a highly concordant pooled estimate of 73.3% (95% CrI 65.3-80.2%), with I2=87.5% and tau=0.833, and satisfactory MCMC convergence (R-hat=1.000).

By outcome domain, pooled successful outcomes were highest for diagnostic pathways (89.3%, 95% CI 70.1-96.7%), followed by timely diagnosis (76.3%, 95% CI 62.9-86.0%), and lowest for diagnostic completion (67.1%, 95% CI 61.8-72.0%) (Qm=5.98, p=0.050). Timely diagnosis demonstrated particularly high heterogeneity (I2=91.2%), whereas diagnostic completion showed moderate heterogeneity (I2=40.6%).

Across determinant domains, frequentist pooled estimates were 79.5% for child developmental/neurobehavioral factors, 74.2% for family/socioeconomic/perceptual factors, 68.0% for intervention/care-navigation factors, and 63.6% for provider/clinical recognition factors. Bayesian estimates were 76.7% (BF=53.76), 72.9% (BF=226.32), 64.3% (BF=25.60), and 53.7% (BF=0.684), respectively.

Meta-regression indicated that determinant category (Qm=13.48, p=0.004) and effect measure (Qm=7.81, p=0.020) significantly explained between-study variation, whereas age group (p=0.203) and geographic region (p=0.453) did not. Family/socioeconomic factors had significantly larger effect sizes (B=2.703, 95% CI 0.661-4.744; p=0.009), as did child developmental/neurobehavioral factors (B=1.516, 95% CI 0.047-2.985; p=0.043).

Potential small-study effects were detected by two of three asymmetry tests, although the Rosenthal fail-safe N was 1,723. Trim-and-fill identified seven potentially missing estimates, with an adjusted pooled effect of 68.4% (95% CI 27.7-109.1%). Importantly, exclusion of two influential outlying estimates produced a pooled outcome of 77.1% (95% CI 71.6-81.9%), indicating that the principal finding was robust.

**Conclusions:** Approximately three-quarters of observed ASD diagnostic outcomes represented successful access, but the substantial heterogeneity indicates that diagnostic access is highly context-dependent. Families were more likely to successfully navigate diagnostic pathways than to complete diagnostic assessment, while timely diagnosis showed the greatest variability across settings. Family and socioeconomic circumstances and child developmental characteristics emerged as particularly important determinants, whereas provider-related effects were more heterogeneous and uncertain. Improving equitable ASD diagnosis requires interventions spanning the entire diagnostic pathway, including developmental surveillance, screening, referral coordination, family navigation, provider capacity, specialist availability, and mechanisms to ensure completion of diagnostic assessment. Greater longitudinal and implementation research is particularly needed in low- and middle-income countries, where diagnostic infrastructure and specialist capacity remain limited.

## 1.0 Introduction

Autism spectrum disorder (ASD) is a lifelong neurodevelopmental condition characterized by persistent differences in social communication and interaction alongside restricted, repetitive patterns of behavior, interests, or activities. The global prevalence of ASD has increased substantially over the past two decades, reflecting improvements in awareness, screening practices, diagnostic criteria, and access to assessment services rather than changes in disease occurrence alone. Despite these advances, access to timely and equitable diagnostic services remains a major public health challenge worldwide. Recent evidence suggests that although signs of ASD can be reliably identified during the second year of life, the average age at diagnosis in many settings remains approximately 4–5 years, with considerable variation across and within countries depending on health system capacity, socioeconomic context, and availability of specialized diagnostic services^1,2^.

Timely diagnosis represents a critical entry point into evidence-based interventions, educational support, family counseling, and community services. Earlier identification has consistently been associated with earlier initiation of intervention, improved developmental and adaptive outcomes, better family preparedness, and more efficient care planning. Conversely, delayed diagnosis may result in missed opportunities for early intervention, prolonged caregiver uncertainty, increased financial burden, and widening health inequities, particularly among underserved populations and low-resource settings. Consequently, improving access to timely diagnostic services has become a global priority in child health and neurodevelopmental care^3^.

Recognizing the importance of early identification, many countries have strengthened developmental surveillance, autism screening, referral pathways, and multidisciplinary diagnostic services over the past decade. International clinical guidelines continue to recommend developmental monitoring throughout early childhood, prompt evaluation of developmental concerns, and timely referral for comprehensive ASD assessment. Nevertheless, substantial disparities persist in access to diagnostic services between countries and across regions within countries. Long waiting times, shortages of trained specialists, fragmented referral systems, socioeconomic disadvantage, limited caregiver awareness, stigma, and differences in healthcare organization continue to impede timely diagnosis, particularly in low- and middle-income countries. These disparities highlight that access to ASD diagnosis is influenced not only by clinical characteristics of the child but also by broader family, provider, and health-system factors^3,4^.

Growing evidence demonstrates that determinants of access to ASD diagnostic services operate across multiple levels of the healthcare pathway. Child-level characteristics such as developmental delay, intellectual disability, language impairment, or co-occurring neurodevelopmental and psychiatric conditions may influence recognition of autism symptoms and referral for diagnostic assessment. Family-level factors—including parental knowledge, health-seeking behavior, socioeconomic status, and stigma—can affect care-seeking and engagement with diagnostic services. Similarly, provider-related determinants, including developmental surveillance practices, responsiveness to parental concerns, use of validated screening tools, and referral decisions, play a crucial role in facilitating or delaying diagnostic completion. Health-system interventions such as family navigation programs have also demonstrated promise in reducing barriers and improving completion of diagnostic evaluations among children at risk for ASD^5^.

Several systematic reviews have examined specific aspects of ASD diagnosis. A recent systematic review identified clinical, social, and environmental predictors associated with earlier diagnosis but synthesized studies narratively because of substantial methodological heterogeneity. Likewise, a systematic review and meta-analysis estimated the global age at ASD diagnosis, while a more recent review highlighted considerable inconsistency in how delayed diagnosis is defined across the literature. Collectively, these reviews have advanced understanding of diagnostic timing but have not comprehensively quantified the determinants influencing access to diagnostic services across the continuum from referral and diagnostic completion to diagnostic pathways and timely diagnosis^5,6^.

To our knowledge, no previous systematic review has synthesized and quantitatively pooled evidence across the broad range of clinical, family, provider, and health-system determinants associated with access to ASD diagnostic services. Understanding these determinants is essential for informing interventions that reduce diagnostic inequities, strengthen referral pathways, improve diagnostic completion, and support earlier identification of autistic children across diverse healthcare settings. Therefore, the objectives of this systematic review and meta-analysis were to: (1) systematically identify and synthesize quantitative evidence on factors associated with access to ASD diagnostic services; (2) evaluate determinants of diagnostic completion, diagnostic pathways, and timely diagnosis across healthcare systems; (3) quantitatively pool comparable effect estimates through meta-analysis where appropriate; and (4) explore potential sources of heterogeneity using subgroup analyses and meta-regression where sufficient data were available..

## 2.0 Methods

### 2.1 Study design and protocol registration

A systematic review and meta-analysis of determinants of access to autism spectrum disorder (ASD) diagnostic services was conducted and reported in accordance with the Preferred Reporting Items for Systematic Reviews and Meta-Analyses (PRISMA) 2020 statement^7,8^. The review protocol was prospectively registered with the International Prospective Register of Systematic Reviews (PROSPERO: CRD420261326047).

### 2.2 Search strategy and study selection

Our systematic review and meta-analysis included studies published between January 1, 2004, and December 30, 2024, that evaluated determinants of access to autism spectrum disorder (ASD) diagnostic services. Eligible studies included individuals of any age undergoing or having received ASD diagnostic assessment, as well as caregivers or families when outcomes related to diagnostic access were reported. Studies were eligible if they reported at least one outcome related to diagnostic access, including diagnostic completion, diagnostic pathways, diagnostic timeliness, age at diagnosis, diagnostic delay, or barriers and facilitators influencing access to diagnostic services. Determinants could operate at individual, child, family, socioeconomic, geographic, provider, healthcare-system, or policy levels. Quantitative, qualitative, and mixed-methods studies were considered, provided that they contained sufficient information to assess determinants of diagnostic access. Studies focusing exclusively on ASD prevalence, etiology, genetics, clinical characteristics, or comorbidities without reporting diagnostic-access outcomes were excluded. Animal studies, laboratory-based studies, editorials, commentaries, conference abstracts without sufficient information, reviews, meta-analyses, and studies unrelated to ASD diagnostic services were also excluded. Studies without sufficient information to determine eligibility or assess relevant outcomes were excluded.

A comprehensive electronic search was conducted **in** MEDLINE/PubMed, Embase, Scopus, and Web of Science, supplemented by searches of grey literature sources. The search strategy combined controlled vocabulary and free-text keywords relating **to** ASD, diagnosis, diagnostic assessment, diagnostic services, healthcare access, diagnostic pathways, diagnostic completion, diagnostic delay, barriers, facilitators, and determinants, using Boolean operators (AND/OR). No language or geographic restrictions were applied. Reference lists of included studies and relevant systematic reviews were also screened to identify additional eligible studies. The complete database-specific search strategies, including search terms, Boolean operators, controlled vocabulary, and date limits, are provided in this *Supplementary Table S1*.

### 2.3 Eligibility criteria

Studies were eligible if they reported at least one outcome related to access to ASD diagnostic services, including diagnostic completion, diagnostic pathways, diagnostic timeliness, age at diagnosis, diagnostic delay, or barriers and facilitators influencing access to diagnostic services. Determinants could operate at individual, child, family, socioeconomic, geographic, provider, healthcare-system, or policy levels. Studies focusing exclusively on ASD prevalence, etiology, genetics, clinical characteristics, or comorbidities without reporting diagnostic-access outcomes were excluded. Animal studies, laboratory-based studies, editorials, commentaries, conference abstracts without sufficient information, reviews, and studies unrelated to ASD diagnostic services were also excluded. Eligibility for quantitative synthesis was more restrictive. Studies were included in the meta-analysis when they reported an effect estimate with a corresponding measure of precision, or sufficient information to calculate one, for determinants associated with diagnostic completion, diagnostic pathways, timely diagnosis, or diagnostic access. Studies that lacked sufficiently comparable outcomes or statistical information for pooling were retained in the narrative synthesis.

### 2.4 Data extraction

Data were extracted independently by two reviewers [JM and KO] using a standardized data extraction form developed before the review. Extracted information included author and publication year, country and geographic region, study design, study period, healthcare setting, sample size, participant characteristics including age, diagnostic outcome, determinant examined, effect measure, effect estimate and 95% confidence interval, variables included in adjusted analyses, and information required for risk-of-bias assessment. For studies contributing to the narrative synthesis, reported barriers and facilitators were additionally extracted and categorized according to the level at which they operated, including child/individual, family, socioeconomic, geographic, healthcare-system, provider, and policy levels.

Where multiple publications appeared to use the same underlying dataset, publications were assessed for overlap to avoid treating the same study population as independent evidence. Studies were counted at the unique-study level, whereas multiple estimates reported within a study were retained where they represented distinct determinants or outcomes and met the criteria for quantitative synthesis. Disagreements between reviewers were resolved through discussion, with consultation of a third reviewer where necessary.

### 2.5 Risk of Bias/Methodological Quality Assessment

Two reviewers independently assessed the methodological quality of included studies using the Mixed Methods Appraisal Tool (MMAT), applying the appropriate criteria according to study design. Disagreements were resolved through discussion or consultation with a third reviewer. The appraisal findings were used to inform interpretation of the evidence and were not used as exclusion criteria.

### 2.6 Synthesis of findings and statistical analysis

Studies that could not be quantitatively combined were synthesized narratively according to determinant domains, including child developmental and neurobehavioral factors; family, socioeconomic and perceptual factors; provider and clinical recognition factors; and intervention and care-navigation factors. Narrative findings were also considered according to three pre- specified diagnostic outcome domains: diagnostic completion, diagnostic pathways, and timely diagnosis.

Where studies reported sufficiently comparable determinants, outcomes, and effect measures, estimates were extracted and quantitatively synthesized. Eligible measures included odds ratios (ORs), risk ratios (RRs), hazard ratios (HRs), prevalence ratios (PRs), incidence rate ratios (IRRs), and other adjusted measures of association with corresponding measures of precision. Where necessary, effect estimates were transformed to the natural logarithmic scale before pooling and subsequently back-transformed for interpretation.

The quantitative synthesis was conducted using JASP ^9,10^. Random-effects models were estimated using restricted maximum likelihood (REML). A random-effects approach was specified a priori because substantial clinical and methodological heterogeneity was anticipated across studies with respect to populations, healthcare settings, determinants, outcome definitions, and study designs.

For the primary synthesis, pooled estimates were expressed on a common probability scale where appropriate to facilitate interpretation across the included effect estimates. The resulting pooled estimates therefore represent standardized summary estimates of successful diagnostic outcomes rather than a single directly measured probability across all studies. Between-study heterogeneity was assessed using Cochran’s Q statistic, I^2^, and τ^2^. Ninety-five percent prediction intervals were calculated to describe the expected range of outcomes in comparable future settings.

Influence diagnostics and Baujat plots were examined to identify studies or effect estimates contributing disproportionately to the pooled estimate or observed heterogeneity. Funnel plots and residual funnel plots were examined where sufficient studies were available to assess potential small-study effects and funnel plot asymmetry.

### 2.7 Subgroup analyses

Pre-specified subgroup analyses were conducted according to outcome domain, effect measure, geographic region, and determinant category. Outcome-domain analyses classified effects into three domains: diagnostic completion, representing outcomes reflecting successful completion or ascertainment of an ASD diagnosis; diagnostic pathways, representing outcomes reflecting the pathway, route, or process through which individuals accessed ASD diagnosis; and timely diagnosis, representing outcomes reflecting early, delayed, or timely receipt of an ASD diagnosis.

The subgroup analysis by determinant category classified determinants into four broad domains: Intervention and Care Navigation Factors; Provider and Clinical Recognition Factors; Child Developmental and Neurobehavioral Factors; and Family, Socioeconomic and Perceptual Factors. Omnibus subgroup tests were used to assess whether pooled estimates differed across categories. Within each subgroup, heterogeneity was assessed using Q, I^2^, and τ^2^ statistics. Subgroup findings were interpreted cautiously where categories contained few effect estimates.

### 2.8 Meta-regression analysis

Random-effects meta-regression analyses were conducted to investigate potential sources of between-study heterogeneity. Pre-specified moderators included participant age group, determinant category, effect measure, and geographic region, where sufficient data were available. Omnibus tests were initially performed to determine whether each moderator explained significant variation in effect sizes, followed by examination of individual regression coefficients to identify specific category differences. Multi-collinearity among moderators was assessed using variance inflation factors (VIFs) where appropriate. Moderators represented by sparse or unstable categories were interpreted cautiously, and estimates were not over-interpreted where model support was limited.

### 2.9 Sensitivity and robustness analyses

Sensitivity analyses were conducted to evaluate the robustness of the pooled estimates. These included leave-one-out analyses, influence diagnostics, and exclusion of influential or outlying effect estimates identified through residual and influence analyses. Pooled estimates before and after removal of influential effect estimates were compared to determine whether individual studies materially altered the overall findings. The sensitivity analysis was also used to assess whether the observed heterogeneity and overall conclusions remained stable after exclusion of influential observations.

### 2.10 Bayesian meta-analysis

To complement the frequentist analyses, a Bayesian random-effects meta-analysis was conducted using weakly informative priors. A Normal (0,1) prior was specified for the pooled log-transformed effect, and a Half-Cauchy (0,1) prior was specified for between-study heterogeneity (τ). Posterior pooled estimates, 95% credible intervals, prediction intervals, and between-study heterogeneity were estimated using Markov Chain Monte Carlo (MCMC) sampling.

Model convergence was assessed using R-hat statistics and effective sample sizes (ESS). Posterior distributions were examined to assess the precision and uncertainty of the pooled estimates. Bayesian subgroup analyses were additionally conducted for the determinant categories to complement the frequentist subgroup analyses. Bayes factors (BFs) were used to quantify the strength of evidence for inclusion of each determinant category in the meta-analytic model. Bayesian results were interpreted alongside the frequentist estimates rather than as replacements for them.

### 2.11 Assessment of small-study effects and publication bias

Potential small-study effects were assessed using funnel plots and statistical tests of funnel plot asymmetry where the number of available effect estimates permitted such assessment. Because funnel plot asymmetry can arise from several mechanisms other than publication bias, including genuine between-study heterogeneity and differences in study size or methodology, evidence of asymmetry was interpreted cautiously. A fail-safe N was additionally calculated to assess the extent to which unpublished or missing null findings would be required to alter the overall statistical significance of the pooled result.

## 3.0 Results

### 3.1 Study Selection

#### 3.1.1 Included Articles and Quality Assessment (Systematic Review)

The database search identified 4,899 records. After removing 537 records without associated data, 4,362 records remained for title and abstract screening. Of these, 3,800 records were excluded based on the predefined eligibility criteria, and 562 reports were sought for retrieval. A total of 112 reports could not be retrieved, leaving 450 reports for full-text eligibility assessment. Following full-text evaluation, 428 reports were excluded for predefined reasons, including inappropriate population, study design, determinant or exposure, diagnostic outcome, or insufficient methodological or quantitative information. Ultimately, 22 studies were included in the systematic review, of which 9 contributed to the quantitative meta-analysis, providing 23 effect estimates, while 13 contributed to the narrative synthesis only ^11–32^, met the inclusion criteria and were included in the narrative synthesis. Of the 15 studies included in the narrative synthesis, nine unique studies provided sufficient quantitative data for inclusion in the meta-analysis, contributing 23 quantitative effect estimates. These studies were reported ^11,12,23,26–31^. Several studies contributed multiple effect estimates because they assessed different determinants, diagnostic outcomes, or comparison groups within the same study population. Therefore, the 23 quantitative effect estimates represented nine unique studies and were not treated as 23 independent studies *(Figure 1)*.

**Figure 1.**
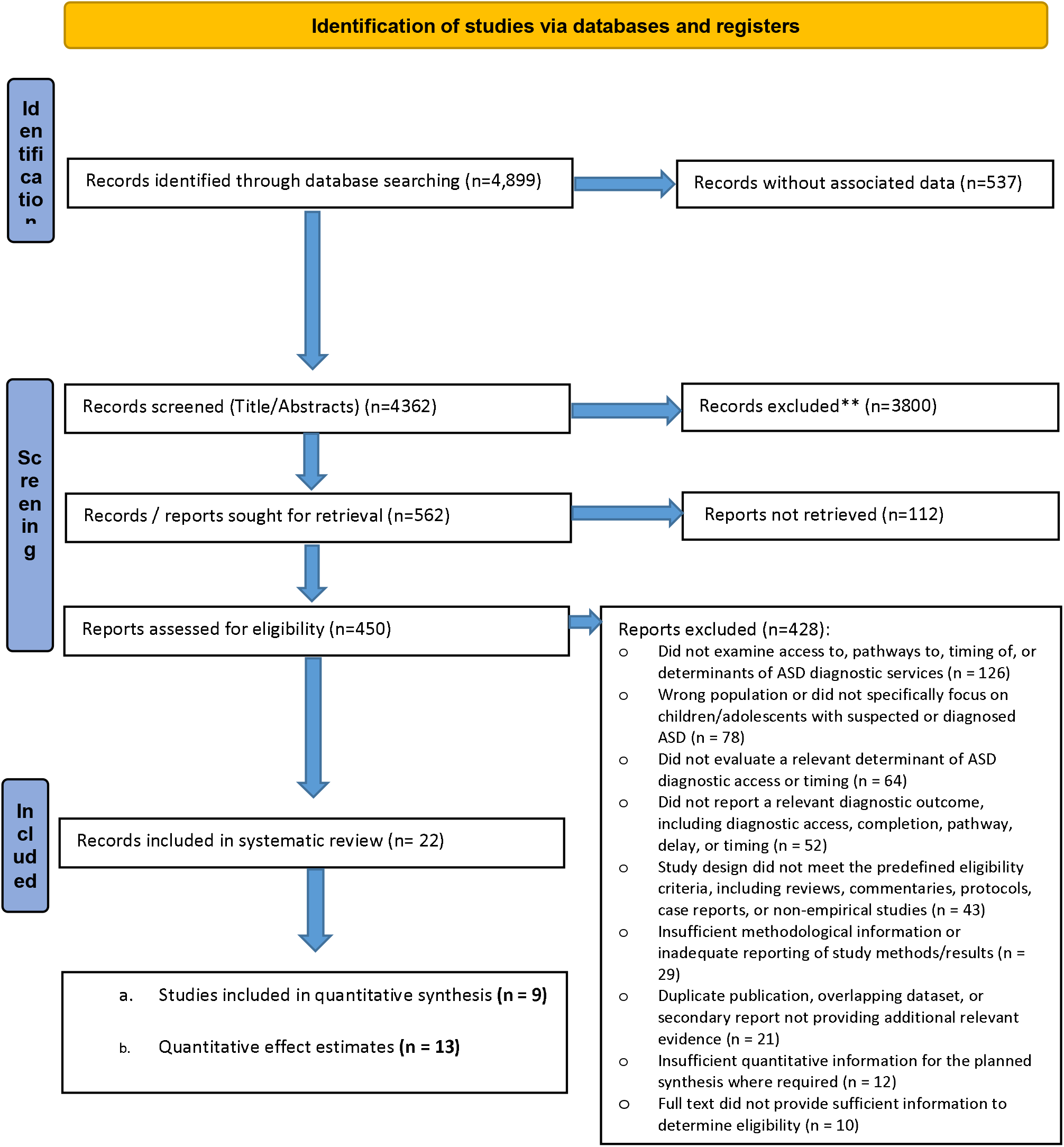
PRISMA flow diagram of study identification, screening, eligibility assessment, and inclusion.

#### 3.1.2 Features of the Included Studies

The 15 included studies ^11–32^ evaluated a broad range of determinants of access to ASD diagnostic services, including socioeconomic, geographic, individual and family, communication, screening, and health-system factors. The studies examined determinants operating across multiple levels, from individual and family characteristics to community, geographic, and health-system factors. Nine unique studies ^11,12,23,26–31^ contributed to the quantitative meta-analysis, providing 23 effect estimates, while thirteen studies ^13–22,24,25,32^ contributed to the narrative synthesis only because their findings were not sufficiently comparable or lacked the statistical information required for quantitative pooling. The quantitative studies were conducted predominantly in the United States and United Kingdom, with additional evidence from Kenya, and reported associations using hazard ratios (HRs)^11,12,23^, adjusted odds ratios (AORs)^27,29,33^, adjusted relative risk ratios (aRRRs)^30^, and adjusted prevalence ratios (APRs)^31^. The studies included a range of observational and other study designs and assessed outcomes related to diagnostic completion, diagnostic pathways, and timely or late diagnosis. Overall, the evidence suggested that socioeconomic disadvantage, rural residence, limited health-system capacity and coordination, communication barriers, and missed or inconsistently implemented screening opportunities may contribute to delayed or inequitable access to ASD diagnostic services *(Table 1)*.

**Table 1.**
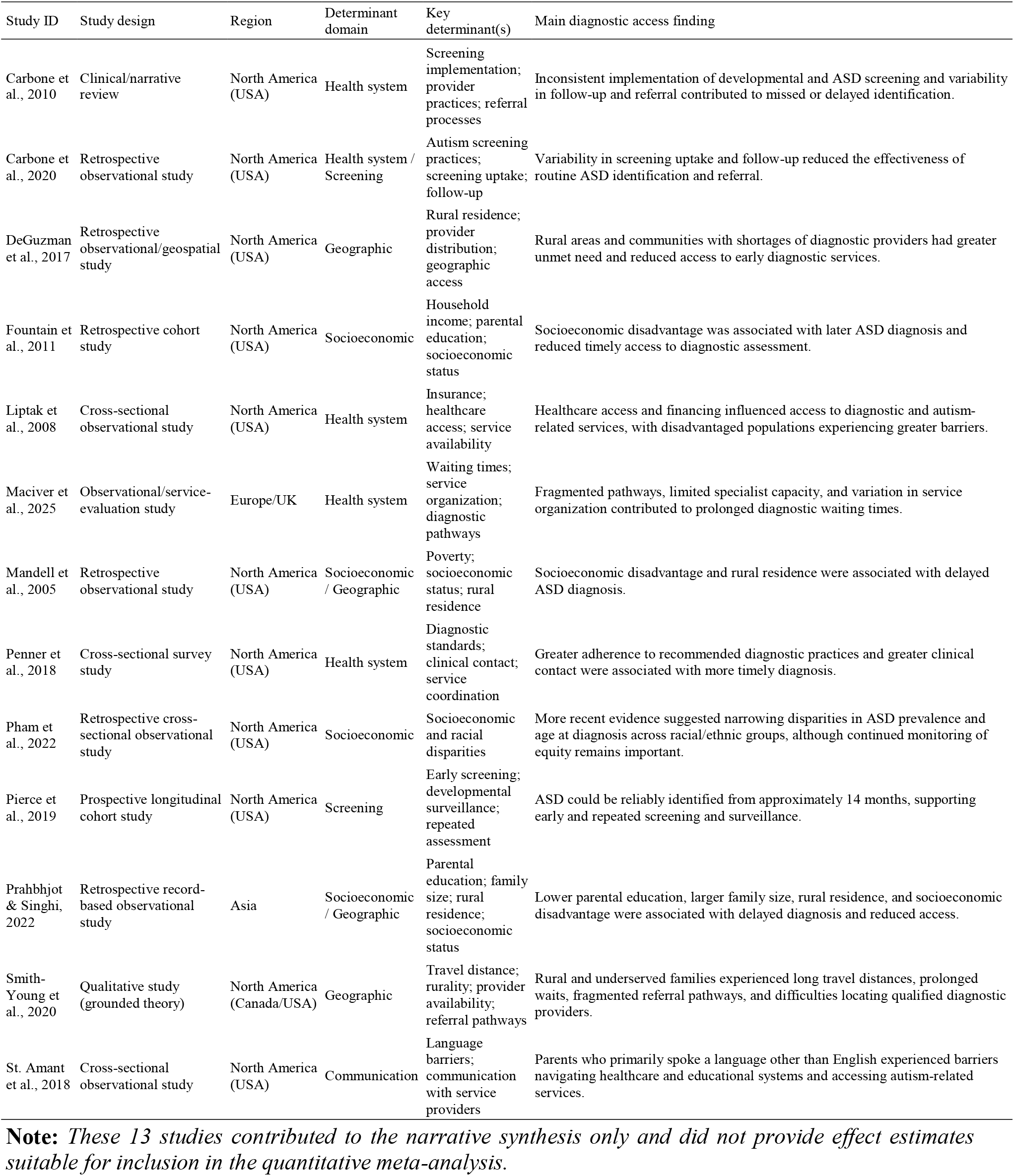
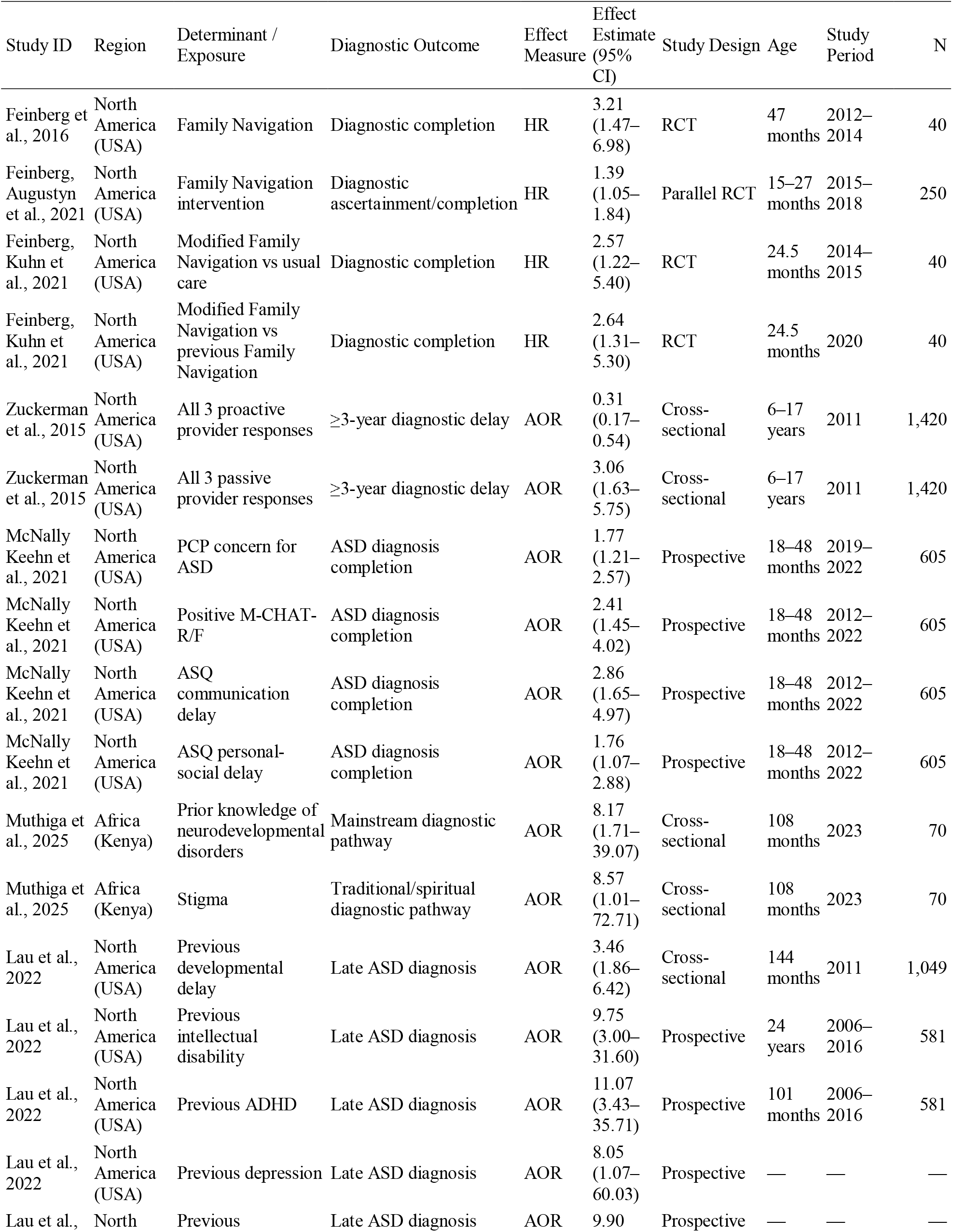
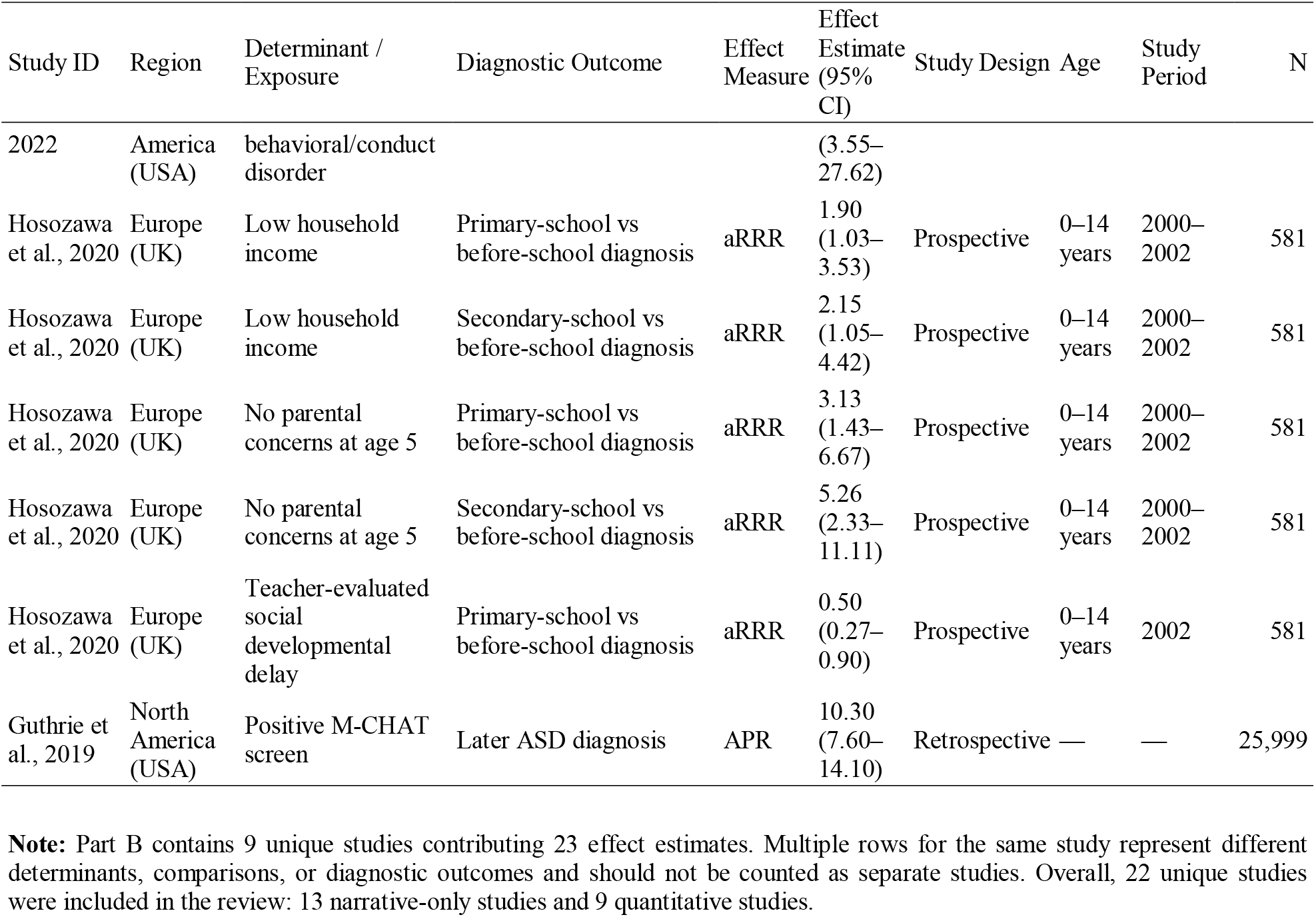
Characteristics and Synthesis of Studies Included in the Systematic Review of Determinants of Access to ASD Diagnostic Services.

#### 3.1.3 Risk of Methodological Quality Assessment

Methodological quality was assessed for all 22 unique studies, comprising 13 studies contributing to the narrative synthesis and 9 contributing to the quantitative synthesis. Studies reporting multiple determinants or effect estimates were assessed once at the study level, so the 23 quantitative effect estimates were not treated as independent studies. The Mixed Methods Appraisal Tool (MMAT), 2018 version^34^, was used, with design-specific criteria applied to randomized, observational, descriptive, and qualitative studies. Two reviewers independently assessed methodological quality, resolving disagreements through discussion or a third reviewer. MMAT assessments informed interpretation of the findings but were not used as exclusion criteria. Detailed study-level assessments are presented (*Table 2*).

**Table 2.**
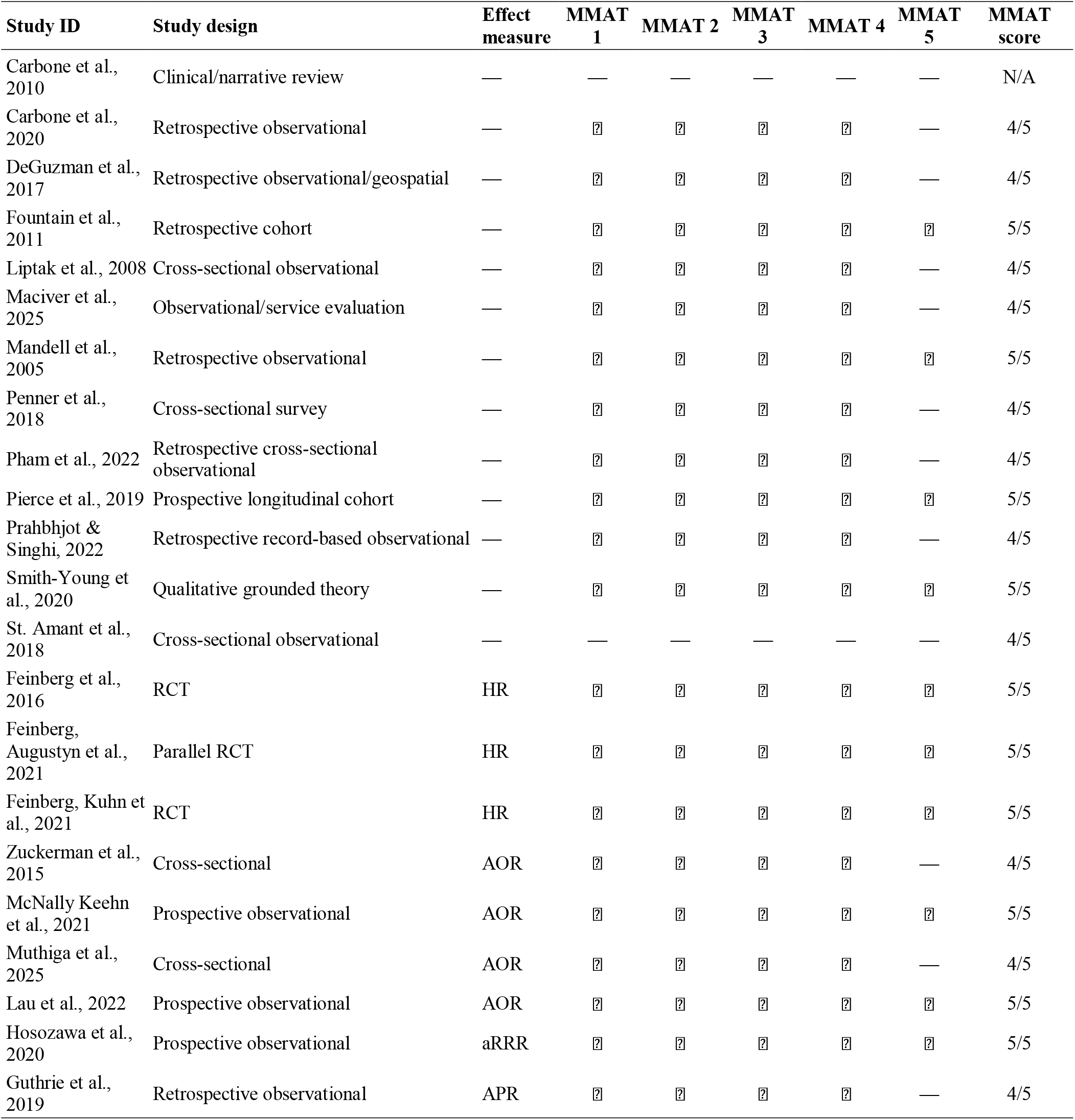
Methodological Quality Assessment of Included Studies Using the Mixed Methods Appraisal Tool (MMAT), 2018.

#### 3.1.4 Socioeconomic Determinants

Five studies examined socioeconomic determinants of access to ASD diagnostic services and consistently showed that socioeconomic disadvantage was associated with delayed diagnosis and reduced access to timely diagnostic services^13–15,30,32^. Lower household income and poverty were repeatedly identified as important predictors of delayed ASD diagnosis, with children from economically disadvantaged families being more likely to receive diagnoses after school entry or during adolescence than children from more affluent households^13,30,32^. Similarly, lower parental educational attainment was associated with later diagnosis, whereas higher maternal or parental education facilitated earlier recognition of developmental concerns and more timely access to diagnostic assessment^15,32^. Geographic residence also interacted with socioeconomic circumstances, as children living in rural communities experienced longer delays in diagnosis than their urban counterparts, reflecting reduced availability of specialist services and diagnostic resources^13,15^. Family characteristics further influenced diagnostic timing, with larger family size associated with delayed diagnosis, particularly in rural settings (4). Although earlier studies reported socioeconomic and racial disparities in ASD diagnosis, more recent evidence suggests that some inequities may be narrowing. A large U.S. cohort study found that by 2021, the prevalence and median age at ASD diagnosis were comparable across White, Black, and Hispanic children, suggesting improvements in screening practices and access to diagnostic services over time, while emphasizing the need for continued monitoring to ensure equitable access across populations^14^. Overall, the evidence indicates that socioeconomic resources—including household income, parental education, and community-level educational and healthcare resources—remain important determinants of timely access to ASD diagnostic services, although recent improvements suggest that some disparities are decreasing in high-income settings.

#### 3.1.5 Geographic Determinants

Three studies examined geographic determinants of access to ASD diagnostic services and consistently identified place of residence and the distribution of diagnostic resources as important influences on timely diagnosis^15–17^. Across the studies, children residing in rural or underserved communities experienced delayed ASD diagnosis and reduced access to specialized diagnostic services compared with those living in urban areas. Rural residence was associated with later diagnosis, reflecting limited availability of specialist providers and healthcare resources^15^. Similarly, geospatial analyses identified rural counties and areas with provider shortages as having substantial unmet needs for early ASD diagnostic services, with geographic location, uneven provider distribution, and insurance-related barriers contributing to delayed diagnostic evaluations^16^. Qualitative evidence further demonstrated that families living in rural or underserved areas frequently encountered long travel distances, fragmented referral pathways, prolonged waiting times, and difficulties locating qualified diagnostic providers. These geographic barriers often interacted with socioeconomic disadvantage, as families with greater financial resources were better able to travel for assessments or seek private diagnostic services, whereas lower-income families experienced additional delays^17^. Overall, the evidence indicates that geographic inequities in the availability and distribution of ASD diagnostic services remain a significant barrier to timely diagnosis, particularly for rural and underserved populations.

#### 3.1.6 Health System Determinants

Six studies examined health system determinants of access to ASD diagnostic services and consistently demonstrated that healthcare organization, service delivery, referral pathways, provider capacity, and adherence to best-practice guidelines substantially influenced timely diagnosis^18,21,22,24,25,30^. Across the studies, prolonged waiting times were primarily attributed to health system factors, including fragmented referral pathways, limited specialist availability, inconsistent diagnostic practices, inadequate care coordination, and variation in service organization across healthcare settings rather than child-level characteristics^22,24^. Greater adherence to recommended diagnostic standards, standardized multidisciplinary assessment pathways, and increased clinical contact were consistently associated with shorter waiting times and more timely diagnostic evaluations^22,24^.

Several studies further highlighted the importance of equitable healthcare delivery in facilitating diagnostic access. Limited healthcare resources, inadequate provider training, insufficient consultation time, and poor coordination within primary care delayed recognition of developmental concerns and hindered timely referral for specialist assessment^21^. Similarly, disparities in access to healthcare services disproportionately affected underserved populations, while enrollment in public insurance programs improved access to diagnostic and autism-related services, emphasizing the role of healthcare financing and equitable service provision in reducing diagnostic delays^25^.

Evidence also demonstrated that the effectiveness of early ASD identification depends not only on the availability of screening tools but also on their consistent implementation within healthcare systems. Inconsistent uptake of routine autism screening, omission of recommended follow-up procedures, and variability in referral practices reduced the effectiveness of primary care screening programs and contributed to missed or delayed diagnoses^21^. Likewise, opportunities for early identification were frequently missed despite the presence of developmental concerns recognized by parents or teachers, suggesting deficiencies in referral pathways and coordination between healthcare and educational services^30^. Overall, the evidence indicates that strengthening healthcare system capacity, improving coordination of multidisciplinary services, standardizing diagnostic pathways, expanding specialist availability, and ensuring timely referral following developmental concerns or positive screening are critical health system strategies for improving access to timely ASD diagnosis.

#### 3.1.7 Communication Determinant

Two studies examined communication-related determinants of access to ASD diagnostic services and consistently demonstrated that effective communication between families, healthcare providers, and educational professionals facilitates timely diagnosis, whereas communication barriers contribute to delays in accessing diagnostic and support services^20,30^. Language barriers were identified as an important obstacle to navigating healthcare and educational systems, with children whose parents primarily spoke a language other than English receiving fewer autism-related support services and having reduced access to appropriate educational and disability resources, even after adjustment for demographic factors^20^. Similarly, effective communication of developmental concerns between parents, teachers, and healthcare providers was critical for timely referral and diagnosis. Children whose parents expressed early concerns about their social development were more likely to receive an earlier ASD diagnosis, whereas the absence of initial parental concerns and missed opportunities to act upon concerns raised by parents or teachers contributed to delayed diagnosis, often despite socio-behavioral difficulties being evident from an early age^30^. Overall, the evidence suggests that both linguistic accessibility and effective communication across families, educational settings, and healthcare services are essential determinants of equitable and timely access to ASD diagnostic services.

#### 3.1.8 Screening Determinants

Three studies examined screening-related determinants of access to ASD diagnostic services and consistently demonstrated that early, systematic, and well-implemented screening facilitates timely diagnosis, whereas missed screening opportunities and inconsistent implementation contribute to diagnostic delays^18,19,27^.Across the studies, early developmental surveillance and routine autism screening enabled earlier identification of children with ASD and promoted more timely referral for comprehensive diagnostic assessment. However, opportunities for early identification were frequently missed despite observable developmental concerns, particularly among children with typical-range intelligence and those from socioeconomically disadvantaged households, suggesting that existing screening and surveillance processes may be less effective for some populations^27^. The effectiveness of universal screening also depended on high screening uptake, adherence to recommended screening protocols, and timely referral following positive screening results. Inconsistent screening practices, lower screening rates among certain populations and healthcare providers, omission of recommended follow-up assessments, and variability in referral practices reduced the effectiveness of screening programs and contributed to missed or delayed diagnoses^18^. Furthermore, evidence from a prospective cohort study demonstrated that ASD can be diagnosed reliably from approximately 14 months of age, supporting the value of screening before the currently recommended 18-month age while highlighting that a single screening assessment may fail to identify a substantial proportion of children who later meet diagnostic criteria, thereby reinforcing the need for repeated developmental surveillance and ongoing re-evaluation^19^. Overall, the evidence indicates that effective implementation of universal screening programs, combined with repeated developmental surveillance and timely referral pathways, is essential for improving early identification and reducing delays in ASD diagnosis.

### 3.2 Pooled Effect and Heterogeneity of Determinants of Access to ASD Diagnostic Services

The quantitative synthesis included nine unique studies from the United States, United Kingdom, and Kenya, contributing 23 quantitative effect estimates ^11,12,23,26–31^. Because the included studies reported different effect measures, the reported estimates were harmonized using their corresponding effect sizes and standard errors for quantitative synthesis. The random-effects meta-analysis yielded a pooled effect of 74.1% (95% CI: 65.8%–81.1%). The pooled effect was statistically significant (*t* (22) = 5.46, *p* < 0.001). Substantial between-study heterogeneity was observed (Q (22) = 209.95, *p* < 0.001), with a between-study standard deviation of τ = 0.832 (95% CI: 0.587–1.238) and between-study variance of τ^2^ = 0.691 (95% CI: 0.344–1.533). The I^2^ was 88.4% (95% CI: 79.1%–94.4%), indicating that most of the observed variability was attributable to differences between studies rather than sampling error. The 95% prediction interval ranged from 32.8% to 94.4%, indicating substantial variation in the expected diagnostic access outcome across different populations and settings (*Table 3; Figure 3)*.

**Figure 3.**
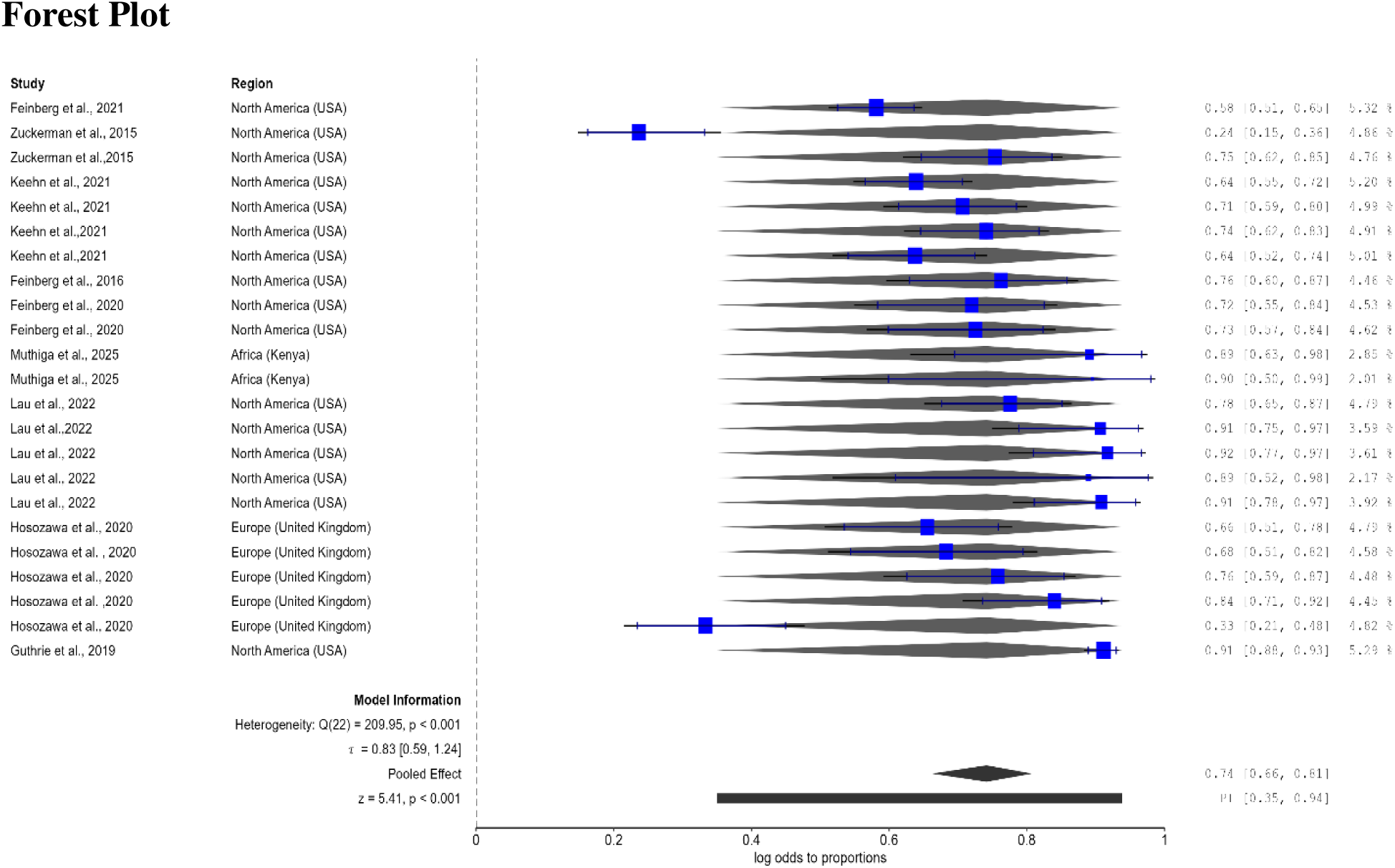
Forest plot of pooled diagnostic access outcome. Random-effects meta-analysis showing a pooled effect of 74.1% (95% CI: 65.8%–81.1%, *p* < 0.001), with substantial heterogeneity (I^2^ = 88.4%).

**Table 3.** Meta-Analytic Summary of Determinants of Access to ASD Diagnostic Services.

| Parameter | Estimate | 95% CI | 95% PI |
| --- | --- | --- | --- |
| Back-transformed pooled proportion | 74.1% | 65.8%–81.1% | 32.8%–94.4% |
| Between-study SD ($\tau$) | 0.832 | 0.587–1.238 | — |
| Between-study variance ($\tau^2$) | 0.691 | 0.344–1.533 | — |
| $I^2$ (Heterogeneity) | 88.4% | 79.1%–94.4% | — |
| $H^2$ (Heterogeneity) | 8.588 | 4.777–17.825 | — |

#### 3.2.1 Stratified Analyses by Outcome Domain

Stratified analyses showed that the pooled proportion of successful diagnostic outcomes differed borderline significantly across the three outcome domains (Q (2) = 5.98, p = .050). Diagnostic pathways had the highest pooled proportion of 0.893 (95% CI: 0.701–0.967), followed by timely diagnosis at 0.763 (95% CI: 0.629–0.860), while diagnostic completion had the lowest pooled proportion at 0.671 (95% CI: 0.618–0.720). In simple terms, people were most likely to successfully navigate a diagnostic pathway, somewhat less likely to receive a timely diagnosis, and least likely to complete the diagnostic process. This suggests that the likelihood of a successful diagnostic outcome may vary depending on which aspect of the diagnostic process is considered. The pooled effects were statistically significant for all three outcome domains: diagnostic completion (z = 6.02, p < .001), timely diagnosis (z = 3.57, p < .001), and diagnostic pathways (z = 3.28, p = .001). Heterogeneity differed substantially across domains. Diagnostic completion showed moderate heterogeneity (I^2^ = 40.6%, Q (7) = 11.13, p = .133), while diagnostic pathways showed no observed heterogeneity (I^2^ = 0%, Q (1) = 0.00, p = .972). In contrast, timely diagnosis demonstrated very high heterogeneity (I^2^ = 91.2%, Q (12) = 172.43, p < .001), indicating considerable variation in findings across studies, particularly for timely diagnosis. The prediction intervals further illustrate this difference. The estimated proportion for diagnostic completion would be expected to fall between 0.561 and 0.765 in a comparable future setting, whereas the prediction interval for timely diagnosis was considerably wider (0.256–0.968), reflecting substantial between-study variability. For diagnostic pathways, the prediction interval was 0.701–0.967, although this estimate should be interpreted cautiously because this subgroup was based on only two effect estimates from the same study. Overall, the stratified analysis suggests that outcome domain may partly explain differences in diagnostic outcomes, with the highest pooled proportion observed for diagnostic pathways and the lowest for diagnostic completion. However, the substantial heterogeneity for timely diagnosis indicates that findings in this domain vary considerably across studies, populations, and/or definitions of timely diagnosis. The borderline subgroup test (p = .050) suggests that these differences are suggestive rather than definitive evidence that outcome domain influences the likelihood of successful access to ASD diagnostic services *(Table 4)*.

**Table 4.** Stratified Pooled Effects and Heterogeneity by Outcome Domain.

| Outcome domain | Pooled proportion | 95% CI | 95% PI | $I^2$ (%) | $Q_{\chi^2}$ | P-value |
| --- | --- | --- | --- | --- | --- | --- |
| Diagnostic completion | 0.671 | 0.618–0.720 | 0.561–0.765 | 40.6 | 11.13 | .133 |
| Timely diagnosis | 0.763 | 0.629–0.860 | 0.256–0.968 | 91.2 | 172.43 | < .001 |
| Diagnostic pathways | 0.893 | 0.701–0.967 | 0.701–0.967 | 0.0 | 0.00 | .972 |
| Test for subgroup differences | — | — | — | — | $Q_{\chi^2}(2) = 5.98$ | .050 |

**Note.** CI = confidence interval; PI = prediction interval; $I^2$ = percentage of variability due to between-study heterogeneity; $Q_{\chi^2}$ = test of residual heterogeneity within subgroup; $Q_{\chi^2}$ = test of differences between subgroups. The pooled effect is transformed from log odds to proportions. Diagnostic pathways should be interpreted cautiously because this subgroup was based on only two effect estimates from the same study.

#### 3.2.2 Bayesian Pooled Meta-Analysis Results and Diagnostics

The Bayesian meta-analysis shows a high pooled diagnostic access outcome, with a pooled mean of 73.3% (95% CI: 65.3%–80.2%) and an identical median, indicating a symmetric posterior distribution. Substantial between-study variability was observed (I^2^ = 87.5%; τ = 0.833), while the prediction interval (32.8%–93.9%) indicates considerable variation in expected diagnostic access across different populations and settings. The MCMC diagnostics indicate reliable estimation, with low MCMC error, large effective sample sizes (ESS = 15,003 for μ; 8,786 for τ), and R-hat values of 1.000, supporting model convergence. Overall, the Bayesian findings closely corroborate the frequentist meta-analysis, which yielded a pooled outcome of 74.1% (95% CI: 65.8%–81.1%), with both approaches indicating substantial heterogeneity across studies *(Table 5; Figure 4)*.

**Figure 4:**
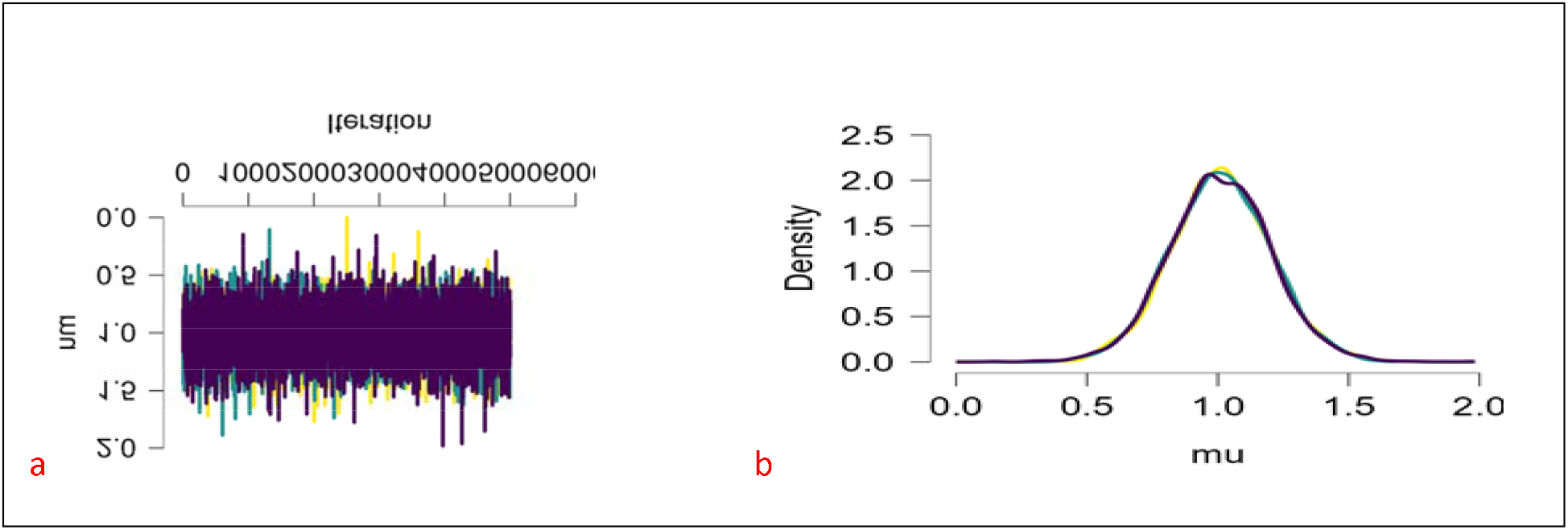
MCMC diagnostic plots for the model parameters: (a) trace plots across multiple chains and (b) autocorrelation function. The trace plots show well-mixed, stable chains, indicating good convergence, while autocorrelation decreases rapidly, indicating efficient sampling and low dependence between successive draws.

**Table 5.** Bayesian Meta-Analytic Estimates for Diagnostic Access.

| Parameter | Estimate | 95% CI | 95% PI / Range |
| --- | --- | --- | --- |
| **Pooled effect ($\mu$, mean)** | 73.3% | [65.3%, 80.2%] | [32.8%, 93.9%] |
| **$\mu$ (median)** | 73.3% | — | — |
| **Between-study heterogeneity** |  |  |  |
| $\tau$ (SD) | 0.833 | [0.579, 1.198] | — |
| $\tau^2$ (variance) | 0.719 | [0.335, 1.436] | — |
| $I^2$ (%) | 87.48 | [78.62, 94.03] | — |
| $H^2$ | 8.884 | [4.678, 16.755] | — |
| **MCMC diagnostics** |  |  |  |
| MCMC error ($\mu$) | 0.002 | — | — |
| MCMC error/SD ($\mu$) | 0.008 | — | — |
| ESS ($\mu$) | 15,003 | — | — |
| R-hat ($\mu$) | 1.000 | — | — |
| MCMC error ($\tau$) | 0.002 | — | — |
| MCMC error/SD ($\tau$) | 0.011 | — | — |
| ESS ($\tau$) | 8,786 | — | — |
| R-hat ($\tau$) | 1.000 | — | — |

**Note.** CI = credible interval; PI = prediction interval; ESS = effective sample size; R-hat = convergence diagnostic. The pooled Bayesian effect estimate (73.3%) closely agrees with the frequentist pooled estimate (74.1%), supporting consistency between the two analytical approaches. Dashes (—) indicate that the measure is not applicable

#### 3.2.3 Assessment of Small-Study Effects and Publication Bias

Funnel plot asymmetry was assessed using three complementary tests. The meta-regression test showed significant asymmetry (*z* = 2.465, *p* = .014), and the rank correlation test was also significant (τ = 0.462, *p* = .002), suggesting possible small-study effects. However, the weighted regression test was not statistically significant (*t* = 0.677, *df* = 21, *p* = .506), providing no evidence of asymmetry by this method. Thus, the findings provide mixed evidence of funnel plot asymmetry, with two of three tests indicating potential small-study effects. The Rosenthal fail-safe N was 1,723, indicating that a large number of additional null studies would be required to reduce the overall finding to non-significance. Overall, the results suggest some evidence of possible small-study effects, but do not provide consistent evidence of publication bias across all tests *(Table 6; Figure 5)*.

**Figure 5.**
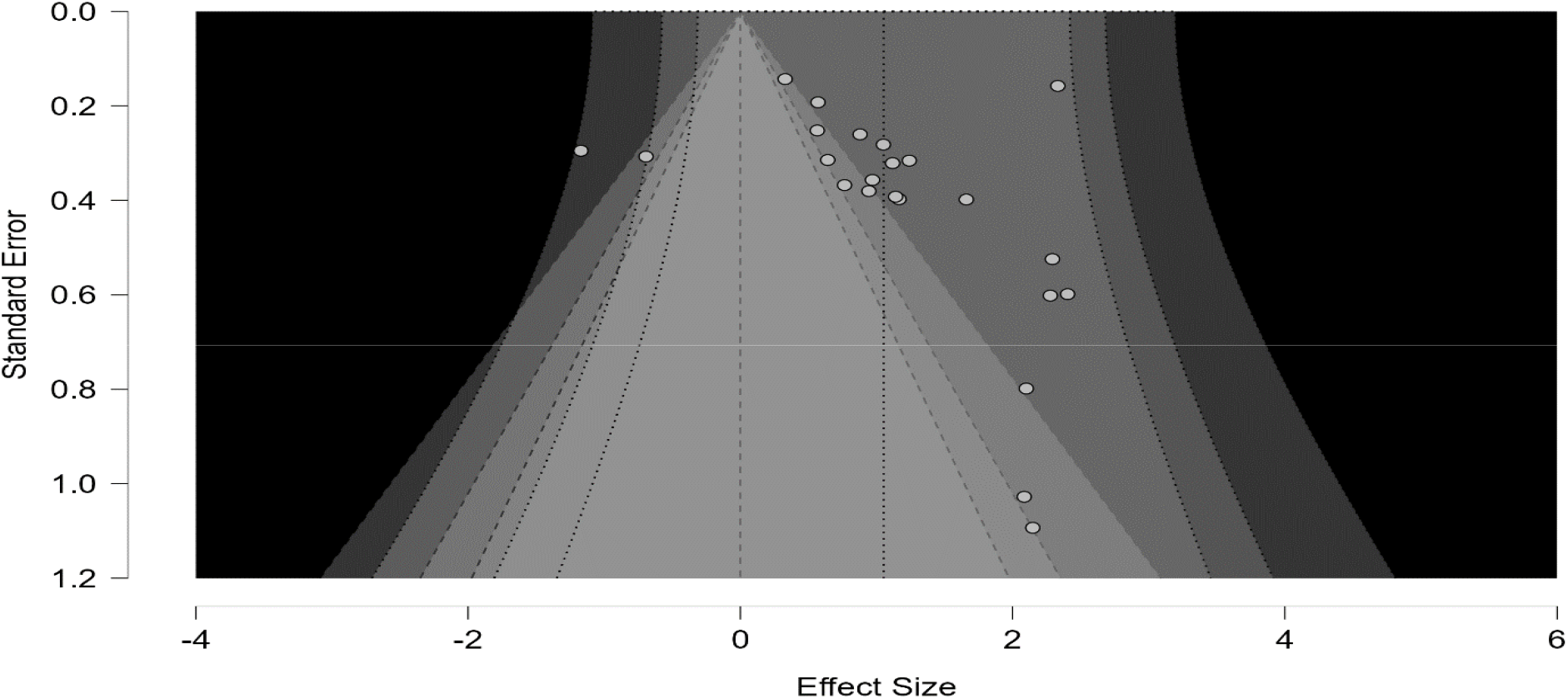
Funnel plot for assessment of small-study effects. Funnel plot showing the distribution of 23 effect estimates around the pooled effect, with evidence of asymmetry suggesting possible small-study effects. Meta-regression (*p* = .014) and rank correlation (*p* = .002) tests supported asymmetry, whereas weighted regression was not significant (*p* = .506).

**Table 6.**

| Method | Statistic | df | p-value | Estimate ($\mu$) | 95% CI |
| --- | --- | --- | --- | --- | --- |
| Meta-regression test | $z = 2.465$ | — | .014 | 0.194 | [-0.567, 0.954] |
| Weighted regression test | $t = 0.677$ | 21 | .506 | 0.651 | [-0.258, 1.560] |
| Rank correlation test | $\tau = 0.462$ | — | .002 | — | — |
| Rosenthal fail-safe N | 1,723 | — | — | — | — |

### 3.3 Assessment of Funnel Plot Asymmetry and the Impact of Trim and Fill Adjustment on Effect Size Estimates

The trim-and-fill analysis identified 7 potentially missing estimates among the 23 effect estimates, producing an adjusted pooled effect of 0.684 (95% CI: 0.277–1.091, *p* < .001) and an adjusted heterogeneity estimate (τ) of 1.024 (95% CI: 0.771–1.491, *p* < .001). The adjusted pooled effect remained statistically significant, indicating that the overall finding persists after accounting for potential missing estimates. However, the wide confidence interval and increased τ suggest considerable uncertainty and substantial between-study variability. Overall, the trim-and-fill results provide evidence that potential small-study effects may influence the observed estimates, but they do not eliminate the evidence of a positive pooled effect *(Table 7; Figure 6)*.

**Figure 6.**
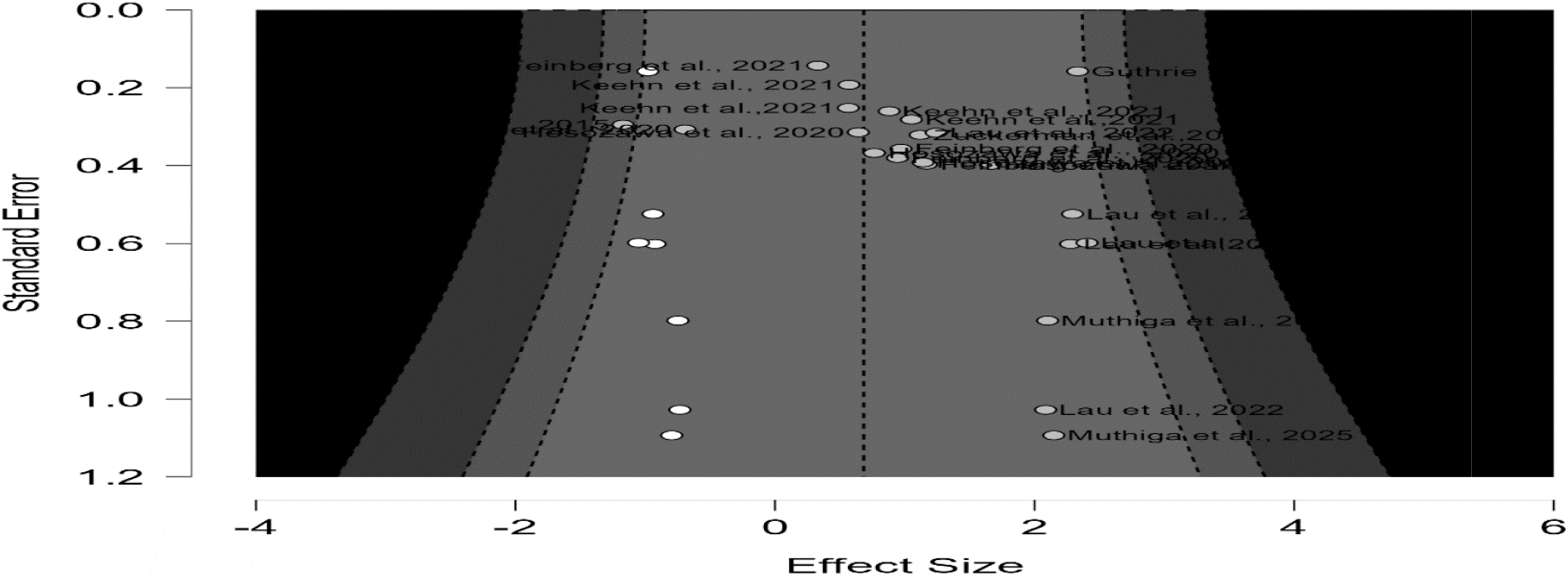
Trim-and-fill analysis showing the impact of potentially missing estimates on the pooled effect. The analysis identified 7 potentially missing estimates and produced an adjusted pooled effect of 0.684 (95% CI: 0.277–1.091, *p* < .001), indicating that the overall effect remained statistically significant after adjustment.

**Table 7.** Trim and Fill Parameter Estimates.

| Parameter | Missing Estimates | Adjusted Estimate | Lower 95% CI | Upper 95% CI | p |
| --- | --- | --- | --- | --- | --- |
| Pooled effect ($\mu$) | 7 | 0.684 | 0.277 | 1.091 | < .001 |
| Heterogeneity ($\tau$) | 7 | 1.024 | 0.771 | 1.491 | < .001 |

**Note.** $\mu$ = pooled effect; $\tau$ = between-study standard deviation; CI = confidence interval. The trim-and-fill procedure identified 7 potentially missing estimates and produced statistically significant adjusted estimates for both the pooled effect and between-study heterogeneity.

### 3.4 Sensitivity Analysis and Robustness of Findings

After conducting a sensitivity analysis involving the removal of two outlier effect estimates from two studies ([Study 1]; [Study 2]), identified from the residual funnel plot (Figure 7a), the overall pooled effect remained robust. The sensitivity analysis yielded a pooled effect estimate of **0.771** (95% CI: 0.716–0.819), corresponding to an estimated probability of a positive outcome of approximately 77% (95% PI: 52.5%–91.1%). Heterogeneity remained statistically significant (Q (20) = 125.41, *p* < .001), with between-study variance of τ^2^ = 0.302 (τ = 0.549). Overall, the findings indicate that removing the two outlier effects had limited impact on the pooled estimate, supporting the robustness of the original findings. However, the remaining prediction interval indicates continued variation in the expected diagnostic access outcome across studies and settings *(Table 8; Figures 7a and 7b)*.

**Figure 7a.**
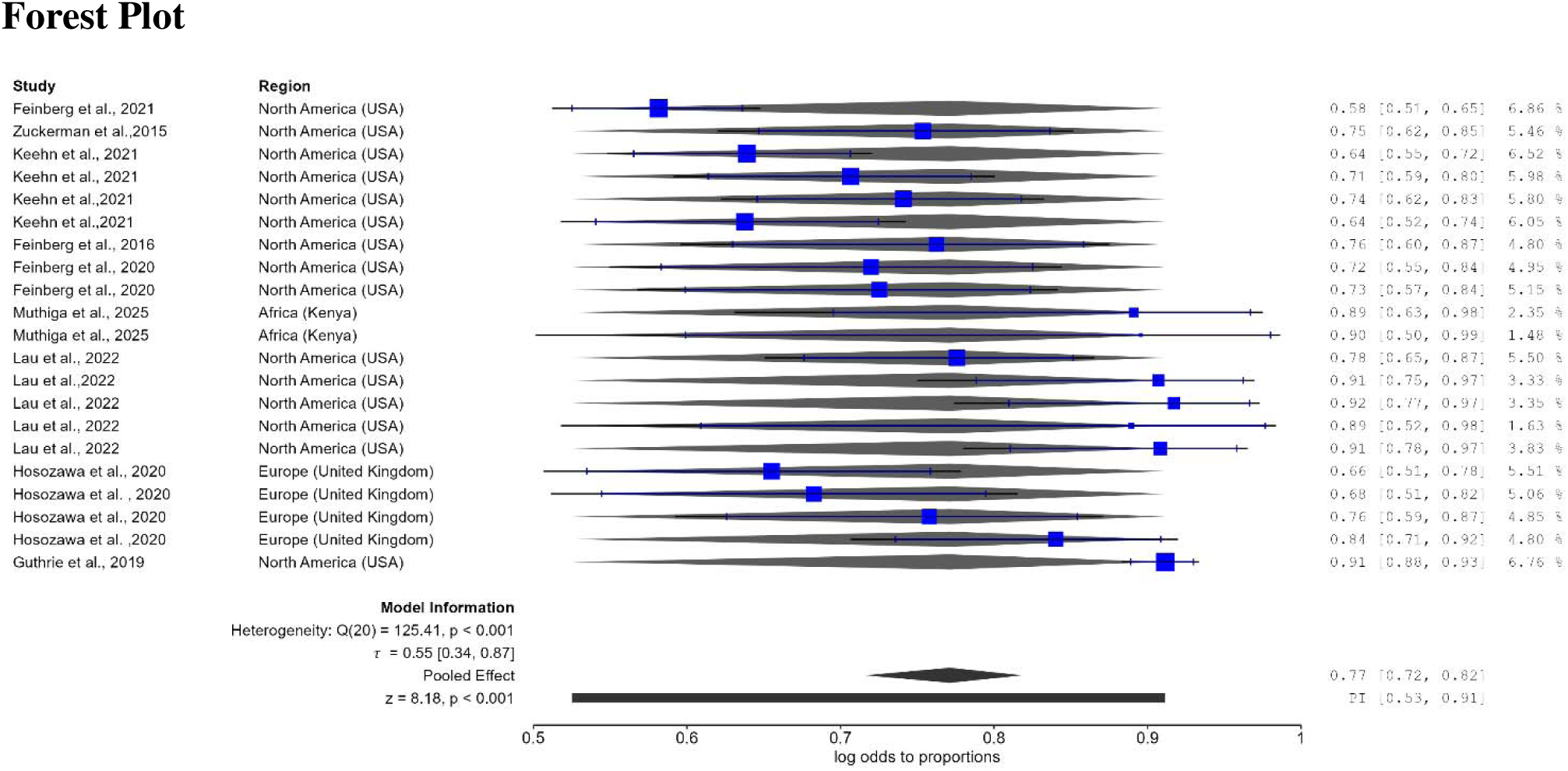
Residual funnel plot identifying outlying effect estimates. Residual funnel plot showing two outlying effect estimates contributed by ^30,33^. These outliers were removed in the sensitivity analysis to assess the robustness of the pooled diagnostic access outcome.

**Figure 7b.**
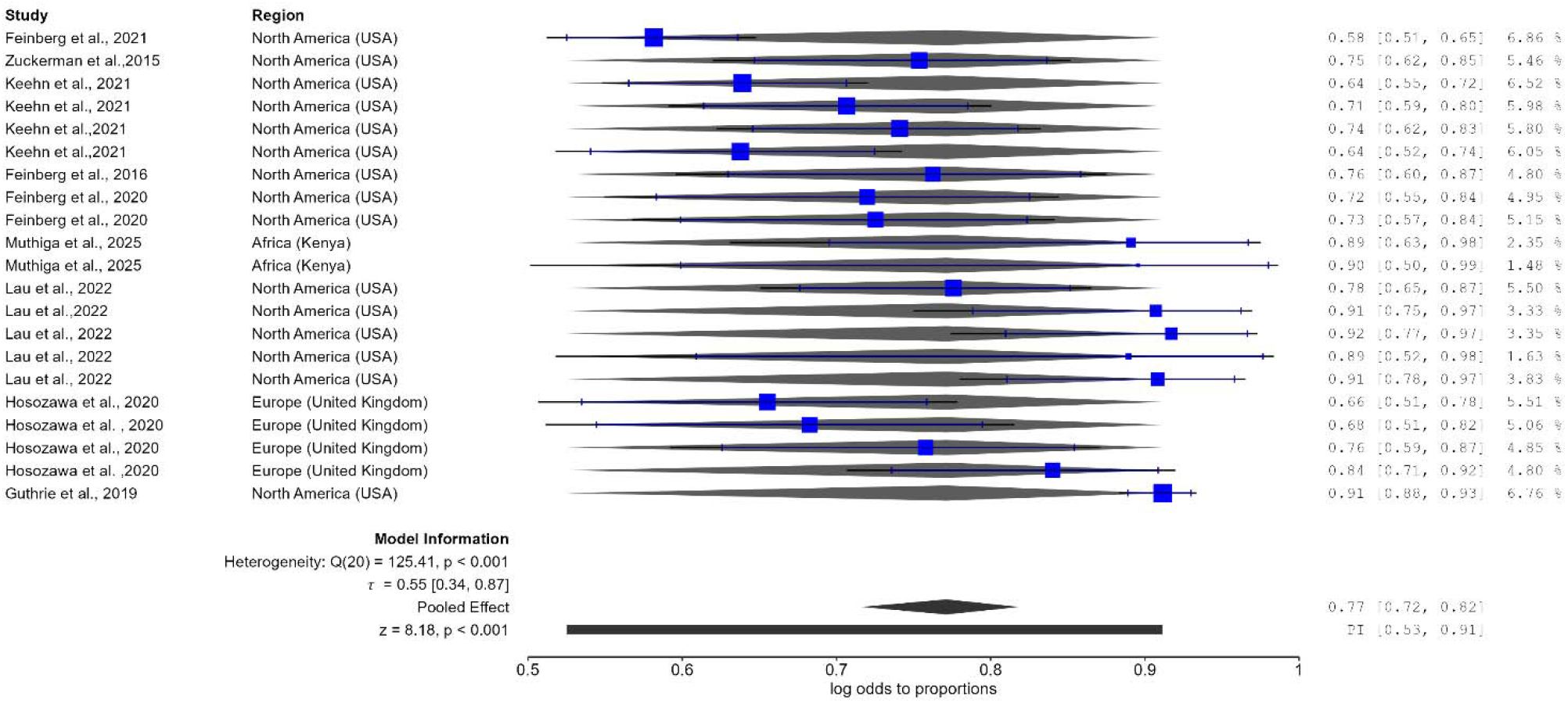
Forest plot of sensitivity analysis after removal of outlier effects. Forest plot showing the pooled diagnostic access outcome after excluding outlier effects from ^30,33^, with a pooled effect of 77.1% (95% CI: 71.6%–81.9%) and substantial residual heterogeneity.

**Table 8.** Sensitivity Analysis of Pooled Diagnostic Access Outcome (Outlier Effects Removed)

| Statistic | Value | 95% CI | 95% PI |
| --- | --- | --- | --- |
| Pooled Effect (logit) | 0.771 | 0.716–0.819 | 0.525–0.911 |
| $\tau$ (between-study SD) | 0.549 | 0.343–0.869 | — |
| $\tau^2$ (between-study variance) | 0.302 | 0.118–0.754 | — |
| $Q\chi^2$ (df = 20) | 125.41 | — | — |
| p-value ($Q\chi^2$) | < .001 | — | — |
| Pooled Effect z | 8.18 | — | — |
| p-value (z) | < .001 | — | — |

**Note.** Two outlier effects from two studies<sup>30,33</sup> were removed. 95% CI = confidence interval; 95% PI = prediction interval; $\tau$ = between-study standard deviation; $\tau^2$ = between-study variance.

### 3.5 Frequentist Subgroup Analysis

#### a) By Effect Measure

The subgroup analysis showed statistically significant pooled effects for HR and AOR, but not for aRRR, with no evidence that the pooled effects differed significantly across effect-measure subgroups. Studies reporting HRs yielded a pooled estimate of 0.680 (95% CI: 0.576–0.768; 95% PI: 0.490–0.824; *p* < .001), with low and non-significant heterogeneity, Q (3) = 7.23, *p* = .065, although the prediction interval indicates some variation across settings. Studies reporting AORs yielded a pooled estimate of 0.768 (95% CI: 0.650–0.855; 95% PI: 0.327–0.957; *p* < .001), but showed substantial heterogeneity, Q (12) = 77.80, *p* < .001, with τ = 0.933 and τ^2^ = 0.871. Studies reporting aRRRs yielded a pooled estimate of 0.664 (95% CI: 0.476–0.811; 95% PI: 0.251–0.921), but the pooled effect was not statistically significant (*z* = 1.71, *p* = .087), with substantial heterogeneity, Q (4) = 26.85, *p* < .001, τ = 0.813 and τ^2^ = 0.660. The APR subgroup could not be pooled because fewer than two estimates were available. Importantly, the test for subgroup differences was not statistically significant, Q (2) = 1.71, *p* = .425, indicating no evidence that the pooled effects differed systematically according to effect measure. Overall, the subgroup findings demonstrate statistically significant effects for HR and AOR, while aRRR did not reach statistical significance, with considerable heterogeneity particularly among AOR and aRRR estimates *(Table 9; Figure 8)*.

**Figure 8.**
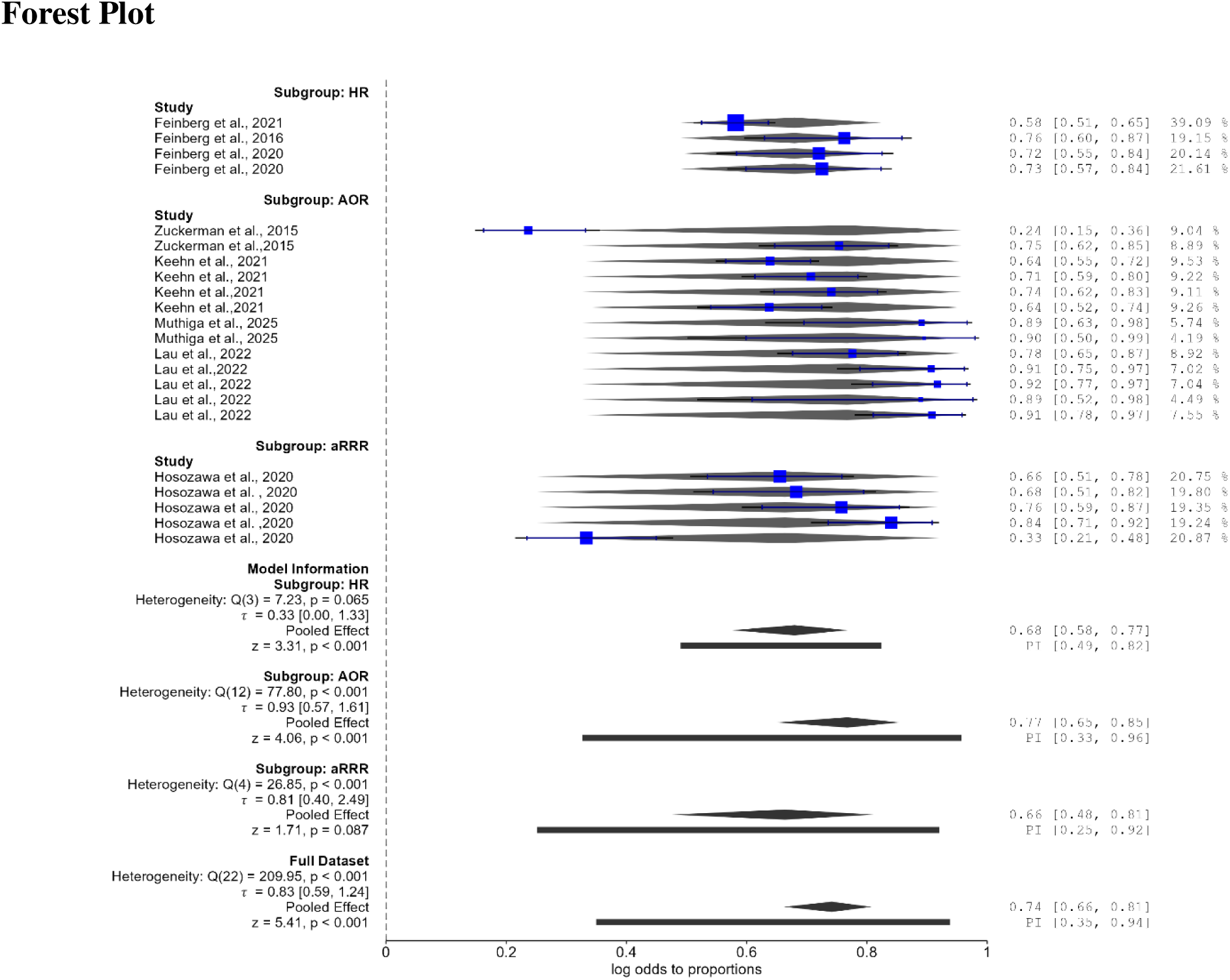
Forest plot of subgroup analysis by effect measure. Forest plot showing pooled effects for HR, AOR, and aRRR estimates, with statistically significant pooled effects for HR and AOR but not aRRR. No significant differences were observed between effect-measure subgroups (*Q* (2) = 1.71, *p* = .425).

**Table 9.** Subgroup Meta-Analytic Effect Estimates by Effect Measure.

| Effect measure | Pooled estimate | 95% CI | 95% PI | P-value | Heterogeneity |
| --- | --- | --- | --- | --- | --- |
| HR | 0.680 | [0.576, 0.768] | [0.490, 0.824] | $< .001$ | $Q^2(3) = 7.23$, $p = .065$ |
| AOR | 0.768 | [0.650, 0.855] | [0.327, 0.957] | $< .001$ | $Q^2(12) = 77.80$, $p < .001$ |
| aRRR | 0.664 | [0.476, 0.811] | [0.251, 0.921] | $.087$ | $Q^2(4) = 26.85$, $p < .001$ |
| APR | — | — | — | — | $< 2$ estimates |
| Subgroup differences | $Q^2(2) = 1.71$ | — | — | $.425$ | — |

**Note.** HR = hazard ratio; AOR = adjusted odds ratio; aRRR = adjusted relative risk ratio; APR = adjusted prevalence ratio; CI = confidence interval; PI = prediction interval. Effect estimates are retained on their original effect-measure scales because HR, AOR, aRRR, and APR are not directly comparable as proportions. The APR subgroup could not be estimated because fewer than two estimates were available. Subgroup differences were not statistically significant, $Q^2(2) = 1.71$ , $p = .425$ .

#### b) By Geographic Region

The subgroup analysis showed variation in the pooled diagnostic access outcome across geographic regions, although the differences between regions were not statistically significant. North America (USA) had a pooled effect of 75.1% (95% CI: 65.6%–82.7%; 95% PI: 34.9%–94.4%; *p* < .001), with substantial between-study heterogeneity, Q (15) = 173.64, *p* < .001, τ = 0.851 and τ^2^ = 0.724. Africa (Kenya) showed the highest pooled effect at 89.3% (95% CI: 70.1%–96.7%; 95% PI: 70.1%–96.7%; *p* = .001), with no observed between-study heterogeneity (τ = 0.000; τ^2^ = 0.000), although the subgroup comprised only two estimates. Europe (United Kingdom) had a pooled effect of 66.4% (95% CI: 47.6%–81.1%; 95% PI: 25.1%–92.1%; *p* = .087) and substantial heterogeneity, Q (4) = 26.85, *p* < .001, τ = 0.813 and τ^2^ = 0.660. The test for subgroup differences was not statistically significant, Q (2) = 3.61, *p* = .165, indicating that geographic region did not significantly explain differences in pooled diagnostic access outcomes. Overall, although the pooled outcome was highest in Kenya and lowest in the United Kingdom, the wide prediction intervals and substantial heterogeneity in the USA and UK indicate considerable variability within regions *(Table 10)*.

**Table 10.** Subgroup Meta-Analytic Effect Estimates by Geographic Region.

| Geographic region | Pooled effect | 95% CI | 95% PI | p-value | Heterogeneity |
| --- | --- | --- | --- | --- | --- |
| North America (USA) | 75.1% | [65.6%, 82.7%] | [34.9%, 94.4%] | < .001 | $Q_{\chi^2}(15) = 173.64$, $p < .001$; $\tau = 0.851$ |
| Africa (Kenya) | 89.3% | [70.1%, 96.7%] | [70.1%, 96.7%] | .001 | $Q_{\chi^2}(1) = 0.00$, $p = .972$; $\tau = 0.000$ |
| Europe (UK) | 66.4% | [47.6%, 81.1%] | [25.1%, 92.1%] | .087 | $Q_{\chi^2}(4) = 26.85$, $p < .001$; $\tau = 0.813$ |
| Subgroup differences | — | — | — | $Q_{\chi^2}(2) = 3.61$,<br>$p = .165$ | — |

**Note.** CI = confidence interval; PI = prediction interval; $\tau$ = between-study standard deviation. Effect estimates are presented as proportions after log-odds-to-proportion transformation. The Kenya subgroup had no observed heterogeneity, but this estimate should be interpreted cautiously given the small number of contributing estimates.

#### c) Determinant Category

The subgroup analysis showed that three of the four determinant categories were significantly associated with ASD diagnostic access, although the magnitude of effects varied across categories. Intervention and Care Navigation Factors had a pooled effect of 68.0% (95% CI: 57.6%–76.8%; 95% PI: 49.0%–82.4%; *p* < .001), with relatively low and non-significant heterogeneity (Q (3) = 7.23, *p* = .065). Provider and Clinical Recognition Factors had a pooled effect of 63.6% (95% CI: 32.3%–86.4%; 95% PI: 9.9%–96.5%), but this effect was not statistically significant (*p* = .400) and showed substantial heterogeneity (Q (3) = 38.42, *p* < .001; τ = 1.249). Child Developmental and Neurobehavioral Factors demonstrated the highest pooled effect at 79.5% (95% CI: 66.8%–88.2%; 95% PI: 34.5%–96.6%; *p* < .001), with substantial heterogeneity (Q (9) = 106.35, *p* < .001; τ = 0.962). Family, Socioeconomic and Perceptual Factors also showed a significant pooled effect of 74.2% (95% CI: 65.0%–81.7%; 95% PI: 59.1%–85.2%; *p* < .001), with relatively low heterogeneity (Q (4) = 5.67, *p* = .225; τ = 0.272). However, the test for subgroup differences was not statistically significant, Q (3) = 2.80, *p* = .424, indicating that the apparent differences in pooled effects across determinant categories were not statistically distinguishable. Overall, the findings suggest that child developmental/neurobehavioral and family-level factors were associated with the highest pooled diagnostic access outcomes, while provider/clinical recognition factors showed the greatest variability and did not demonstrate a statistically significant pooled association. The significant heterogeneity within the provider and child developmental/neurobehavioral categories indicates that individual determinants within these broad domains may operate differently across populations and settings *(Table 11; Figure 10)*.

**Table 11.** Subgroup Meta-Analytic Effect Estimates by Determinant Category.

| **Determinant category** | **Pooled effect** | **95% CI** | **95% PI** | **p-value** | **Heterogeneity** |
| --- | --- | --- | --- | --- | --- |
| *Intervention and Care Navigation Factors* | 68.0% | [57.6%, 76.8%] | [49.0%, 82.4%] | < .001 | $Q(3) = 7.23, p = .065$ |
| *Provider and Clinical Recognition Factors* | 63.6% | [32.3%, 86.4%] | [9.9%, 96.5%] | .400 | $Q(3) = 38.42, p < .001$ |
| *Child Developmental and Neurobehavioral Factors* | 79.5% | [66.8%, 88.2%] | [34.5%, 96.6%] | < .001 | $Q(9) = 106.35, p < .001$ |
| *Family, Socioeconomic and Perceptual Factors* | 74.2% | [65.0%, 81.7%] | [59.1%, 85.2%] | < .001 | $Q(4) = 5.67, p = .225$ |
| **Subgroup differences** | — | — | — | $Q(3) = 2.80, p = .424$ | |

**Note.** CI = confidence interval; PI = prediction interval; $\tau^2$ = between-study standard deviation; $\tau^2$ = between-study variance. Effect estimates are presented as proportions after log-odds-to-proportion transformation. Although pooled effects differed numerically across determinant categories, the test for subgroup differences was not statistically significant.

### 3.6 Bayesian Subgroup Meta-Analysis by Determinant Category

The Bayesian subgroup meta-analysis demonstrated positive pooled effects across all four determinant categories, although the strength of Bayesian evidence and the degree of between-study heterogeneity varied substantially. Intervention and Care Navigation Factors showed a pooled effect of 64.3% (95% credible interval [CrI]: 50.0%–74.7%; 95% prediction interval [PI]: 43.9%–80.7%), with strong evidence for inclusion (BF = 25.60) and relatively low heterogeneity (I^2^ = 27.5%). Provider and Clinical Recognition Factors showed a pooled effect of 53.7% (95% CrI: 38.9%–76.2%; 95% PI: 7.1%–94.9%), but evidence for inclusion was weak (BF = 0.684), while heterogeneity was very high (I^2^ = 89.5%), indicating considerable uncertainty and variability across studies. Child Developmental and Neurobehavioral Factors demonstrated a high pooled effect of 76.7% (95% CrI: 58.0%–86.1%; 95% PI: 25.8%–96.5%), with strong evidence for inclusion (BF = 53.76) and substantial heterogeneity (I^2^ = 88.9%). Family, Socioeconomic and Perceptual Factors also showed a high pooled effect of 72.9% (95% CrI: 64.2%–80.0%; 95% PI: 60.3%–82.4%), with very strong evidence for inclusion (BF = 226.32) and low heterogeneity (I^2^ = 10.8%). Thus, the Bayesian analysis provides the strongest evidence for Family, Socioeconomic and Perceptual Factors, followed by Child Developmental and Neurobehavioral Factors and Intervention and Care Navigation Factors, whereas evidence for Provider and Clinical Recognition Factors was inconclusive. Importantly, these findings closely correspond with the frequentist analysis: the frequentist pooled effects were 68.0%, 63.6%, 79.5%, and 74.2%, respectively, while the Bayesian estimates were 64.3%, 53.7%, 76.7%, and 72.9%. Both approaches therefore identify child developmental/neurobehavioral and family-related factors as having the highest pooled outcomes, while provider/clinical recognition factors show the greatest uncertainty and heterogeneity. Overall, the Bayesian findings reinforce the frequentist results while providing additional information on the strength of evidence and uncertainty within each determinant category *(Table 12; Figure 9)*.

**Figure 9.**
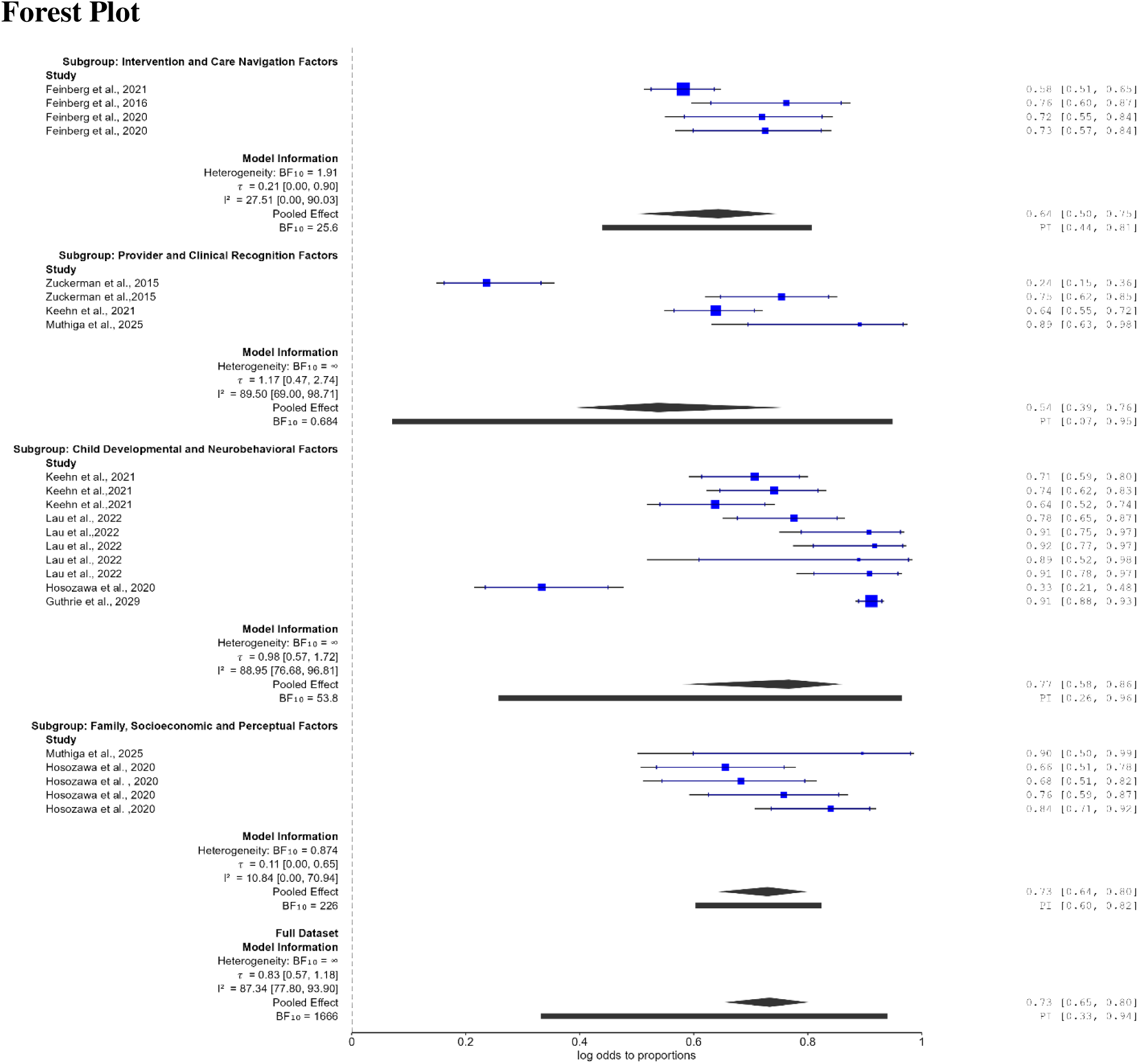
Forest plot of Bayesian subgroup effects by determinant category. Forest plot showing pooled Bayesian effects for Intervention and Care Navigation, Provider and Clinical Recognition, Child Developmental and Neurobehavioral, and Family, Socioeconomic and Perceptual Factors. The highest pooled effects were observed for Child Developmental and Neurobehavioral Factors (0.767) and Family, Socioeconomic and Perceptual Factors (0.729), with corresponding 95% credible and prediction intervals.

**Table 12.** Bayesian Subgroup Meta-Analysis by Determinant Category.

| **Determinant category** | **Effect (Mean)** | **95% CrI** | **95% PI** | **BF** | **I<sup>2</sup> (%)** | **Interpretation** |
| --- | --- | --- | --- | --- | --- | --- |
| Intervention and Care Navigation Factors | 0.643 | 0.500–0.747 | 0.439–0.807 | 25.60 | 27.5 | *Positive effect; strong evidence; low heterogeneity* |
| Provider and Clinical Recognition Factors | 0.537 | 0.389–0.762 | 0.071–0.949 | 0.684 | 89.5 | *Positive but uncertain effect; weak evidence; substantial heterogeneity* |
| Child Developmental and Neurobehavioral Factors | 0.767 | 0.580–0.861 | 0.258–0.965 | 53.76 | 88.9 | *High effect; strong evidence; substantial heterogeneity* |
| Family, Socioeconomic and Perceptual Factors | 0.729 | 0.642–0.800 | 0.603–0.824 | 226.32 | 10.8 | *High effect; very strong evidence; low heterogeneity* |

**Note.** CrI = credible interval; PI = prediction interval; BF = inclusion Bayes factor; I<sup>2</sup> = percentage of total variation attributable to between-study heterogeneity. Effect estimates are presented as proportions after log-odds-to-proportion transformation. Bayes factors >1 support inclusion of the pooled effect, whereas values <1 provide evidence against inclusion. The very large BF for Family, Socioeconomic and Perceptual Factors should be interpreted cautiously because the software notes potential computational instability for very large Bayes factors.

### 3.7 Meta-Regression Analyses

Meta-regression analyses were conducted to examine whether age group, determinant category, effect measure, and region explained variability in effect sizes. The omnibus tests indicated that determinant category significantly moderated effect sizes (Q = 13.483, df = 3, p = .004), as did effect measure (Q = 7.814, df = 2, p = .020). In contrast, neither age group (Q = 1.621, df = 1, p = .203) nor region (Q = 0.563, df = 1, p = .453) significantly moderated effect sizes. Thus, the observed heterogeneity in effect sizes appears to be more strongly related to the type of determinant examined and the statistical effect measure used, rather than differences in participant age group or geographic region.

At the coefficient level, the determinant-category contrasts showed that studies examining Child Developmental and Neurobehavioral Factors had significantly larger effect sizes than the omitted reference determinant category (B = 1.516, 95% CI: 0.047–2.985, p = .043). Similarly, Family, Socioeconomic and Perceptual Factors were associated with significantly larger effect sizes (B = 2.703, 95% CI: 0.661–4.744, p = .009). The contrast for Provider and Clinical Recognition Factors was not statistically significant (B = 0.398, 95% CI: −1.340–2.136, p = .653). For effect measure, the aRRR category was associated with significantly smaller effect sizes relative to the omitted reference effect-measure category (B = −2.597, 95% CI: −4.419 to −0.775, p = .005), whereas the contrast for AOR did not reach statistical significance (B = −1.276, 95% CI: −2.751–0.198, p = .090). The coefficient for age group >60 months was not significant (B = 0.588, 95% CI: −0.317–1.494, p = .203), consistent with the non-significant omnibus age-group test. Likewise, Africa (Kenya) did not differ significantly from the omitted regional reference category (B = 0.683, 95% CI: −1.101–2.467, p = .453). Overall, these findings indicate that determinant category and effect measure were significant sources of between-study variation, whereas age group and region did not provide evidence of systematic moderation of effect sizes *(Table 13)*.

**Table 13.** Meta-regression results examining moderators of effect sizes.

| **Predictor** | **Q$\chi^2$** | **df** | **p-value** |
| --- | --- | --- | --- |
| Age group | 1.621 | 1 | .203 |
| Determinant Category | 13.483 | 3 | **.004** |
| Effect Measure | 7.814 | 2 | **.020** |
| Region | 0.563 | 1 | .453 |

**Note.** Q $\chi^2$ = omnibus test statistic for moderator effects in meta-regression; df = degrees of freedom; p = p-value. Fixed-effects meta-regression tests were conducted using a z-distribution. The model partially removed one coefficient for Effect Measure and one coefficient for Region because of missing values, collinearity, or empty/missing category combinations. Consequently, coefficient-level findings should be interpreted with caution.

### 3.8 Discussion

#### 3.8.1 Review of the Main Findings

This systematic review identified a broad range of individual, family, socioeconomic, geographic, communication, screening, and health-system determinants of access to ASD diagnostic services. The quantitative synthesis of nine unique studies and 23 effect estimates yielded a pooled proportion of 74.1% (95% CI: 65.8%–81.1%), indicating that approximately three-quarters of the observed diagnostic outcomes represented successful access. However, substantial heterogeneity was evident (I^2^ = 88.4%), suggesting that access to ASD diagnosis varies considerably across populations, settings, and diagnostic processes. This finding is consistent with earlier evidence showing that ASD diagnosis is influenced by socioeconomic circumstances, parental recognition, interactions with health and education systems, and geographic availability of services ^35^.

The high heterogeneity is important because access to ASD diagnosis is not a single event but a multistage process involving recognition of developmental concerns, referral, assessment, and completion of diagnosis. Previous reviews similarly describe the pathway as complex, with delays arising from provider knowledge, unclear referral pathways, service shortages, cost, and communication difficulties ^36,37^.Thus, the overall pooled estimate should be interpreted as a summary of a heterogeneous diagnostic process rather than as a universal probability of obtaining an ASD diagnosis.

#### 3.8.1 Outcome Domain and Diagnostic Access

The stratified analysis provides an important refinement of the overall finding. Successful outcomes were highest for diagnostic pathways (89.3%), followed by timely diagnosis (76.3%), and lowest for diagnostic completion (67.1%). In practical terms, families appeared more likely to enter or navigate a diagnostic pathway than to complete the diagnostic process, while timely diagnosis fell between these two outcomes. The borderline subgroup difference (Q = 5.98, p = .050) suggests that the stage or aspect of diagnosis being considered may partly explain variation in access.

This pattern is consistent with qualitative evidence describing ASD diagnosis as a prolonged and sometimes fragmented journey. Parents commonly report difficulty knowing where to seek assessment, navigating multiple services, and dealing with prolonged waiting periods even after concerns have been recognized^38^. The distinction between pathway entry and diagnostic completion is therefore important: recognition and referral do not necessarily translate into completed assessment. The particularly high heterogeneity for timely diagnosis (I^2^ = 91.2%) further suggests that what constitutes a timely diagnosis varies substantially between studies and settings. This is consistent with finding that there is a considerable variation in age at diagnosis and identified socioeconomic status, parental concern, healthcare and education-system interactions, and geographic resources as important contributors^35^.

#### 3.8.3 Socioeconomic, Family and Perceptual Factors

Socioeconomic and family-related factors emerged as an important component of diagnostic access. Lower household income, lower parental education, rural residence, limited resources, and reduced ability to navigate services were repeatedly associated with delayed or less equitable access in the narrative evidence. These findings are consistent with the wider literature showing that socioeconomic advantage and greater parental recognition of developmental concerns are associated with earlier diagnosis ^35^.

The quantitative findings further support the importance of this domain. Family, Socioeconomic and Perceptual Factors had a pooled effect of 74.2%, and the meta-regression showed significantly larger effect sizes for this category relative to the reference category (B = 2.703, p = .009). The Bayesian analysis also provided strong evidence for this determinant category (BF = 226.32) with relatively low heterogeneity. Taken together, these findings suggest that the social and perceptual environment in which families recognize and respond to developmental concerns may be an important determinant of subsequent access.

This interpretation is also supported by qualitative research showing that families experience financial burdens, long waiting periods, difficulties navigating services, and variable responsiveness from healthcare providers during the diagnostic process^38^. Importantly, these barriers may interact rather than operate independently: families with fewer financial resources may have greater difficulty travelling to specialist services, pursuing private assessment, or repeatedly following up referrals.

#### 3.8.4 Child Developmental and Neurobehavioral Factors

Child developmental and neurobehavioral characteristics were associated with the highest pooled outcome in the frequentist analysis (79.5%) and remained strongly supported in the Bayesian analysis (76.7%; BF = 53.76). The meta-regression also indicated significantly larger effect sizes for this category (B = 1.516, p = .043). This finding is plausible because the type, severity, and visibility of developmental and behavioral characteristics can influence whether ASD is recognized, suspected, and referred for diagnostic assessment. Earlier evidence has consistently shown that greater symptom severity, developmental impairment, and greater parental concern are associated with earlier ASD identification and diagnosis ^35^. This association has also been demonstrated in more recent empirical studies. In a Canadian cohort of 421 preschool children, more advanced language and cognitive skills were associated with later diagnosis, while ASD symptom severity contributed independently to variation in age at diagnosis^39^. Similarly, a population-based surveillance study found that children with lower IQ and developmental regression were identified at younger ages, suggesting that more readily observable developmental differences may prompt earlier recognition^40^. More recent evidence from 801 autistic children likewise found that language delay and developmental regression were associated with earlier diagnosis, whereas cognitive and adaptive delays showed more complex relationships with diagnostic timing^41^ Conversely, children with subtler or less readily recognizable presentations may experience longer diagnostic pathways. A study of high-risk children found that those diagnosed later had more advanced language and adaptive skills and milder ASD symptoms than children diagnosed at 18–24 months^42^. Similarly, evidence from a large parent-reported study suggests that clinical signs that are more easily recognized as developmental concerns are more strongly associated with earlier diagnosis, whereas children without early communication deficits may experience delayed diagnosis^5^. Importantly, recent evidence from Kenya also demonstrates that specific developmental and neurobehavioral characteristics—including delayed walking, ADHD, and intellectual developmental disorder—can be associated with diagnostic delay, highlighting the relevance of child-level presentation within lower-resource diagnostic systems ^28^.

The findings also reinforce the importance of systematic developmental surveillance and screening. A study concluded that systematic monitoring and ASD-specific screening during early childhood can facilitate earlier identification, although screening alone does not eliminate downstream barriers to diagnostic assessment^43^. Thus, identifying developmental concerns is only the first step; effective referral and diagnostic capacity are required to convert recognition into completed diagnosis.

#### 3.8.5 Health-System, Provider and Navigation Factors

The review identified health-system capacity, provider recognition, referral coordination, specialist availability, and navigation as important determinants of access. These findings are consistent with broader reviews identifying shortages of services, limited provider awareness, unclear referral pathways, and inadequate coordination as major barriers to ASD healthcare access ^36^.

The evidence concerning Intervention and Care Navigation Factors was particularly relevant. Family navigation was associated with improved access in several quantitative studies, while the pooled estimate for this category was 68.0%. External evidence similarly suggests that navigation can help families overcome practical and system-level barriers. For example, a navigation study among disadvantaged families with children screening positive for ASD found that families commonly experienced multiple barriers to referral and evaluation, with navigation specifically used to address these obstacles^12^.

However, provider and clinical recognition factors showed greater uncertainty. Their pooled effect was not statistically significant in the frequentist analysis, the Bayesian evidence for inclusion was weak (BF = 0.684), and heterogeneity was high. This suggests that provider-related factors may operate differently depending on the healthcare system, provider expertise, clinical presentation, and referral context rather than having a uniform effect across settings.

#### 3.8.6 Geographic and Communication Inequities

Although geographic region did not significantly moderate effect sizes (Q = 0.563, p = .453), the narrative synthesis identified important geographic barriers. Rural and underserved populations may face longer travel distances, fewer specialists, fragmented referral pathways, and longer waiting periods. These barriers are consistent with evidence that geographic variation in diagnostic age reflects differences in community resources and healthcare systems ^13^.

Similarly, communication emerged as an important pathway through which inequities may occur. Effective communication between parents, teachers, primary-care providers, and specialists facilitates recognition and referral, whereas language barriers or dismissal of parental concerns can delay diagnosis. Recent qualitative syntheses have emphasized the importance of healthcare professionals listening to and validating parental concerns and providing clear information throughout the diagnostic process^38^.

#### 3.8.7 Screening and Early Identification

The narrative evidence indicates that screening can improve early identification, but its effectiveness depends on implementation and follow-up. This is consistent with recommendations emphasizing systematic developmental surveillance and ASD screening during early childhood^43^. However, screening should not be interpreted as sufficient on its own. A positive screen must be followed by accessible referral, specialist assessment, and diagnostic completion.

This distinction helps explain why the present review found a relatively high pooled outcome for diagnostic pathways but a lower outcome for diagnostic completion. A health system may successfully identify children at risk and initiate referrals while still failing to provide sufficient capacity to complete diagnostic assessments. Thus, improving screening without strengthening downstream diagnostic capacity may simply shift the point at which families encounter delays.

#### 3.8.8 Interpretation of Heterogeneity and Meta-Regression

The substantial overall heterogeneity (I^2^ = 88.4%) indicates that diagnostic access is highly context-dependent. Importantly, the meta-regression identified determinant category and effect measure, rather than age group or geographic region, as significant moderators. Determinant category therefore appears to capture meaningful differences in the mechanisms through which access is achieved or constrained.

The significant effect-measure moderation should nevertheless be interpreted cautiously. Different measures—HR, AOR, aRRR and APR—represent different statistical quantities and are not necessarily directly interchangeable in substantive meaning. The observed moderation may therefore partly reflect methodological differences between studies rather than true differences in diagnostic access. The partial removal of coefficients for effect measure and region also indicates sparse cells and should further discourage over interpretation of individual contrasts.

#### 3.8.9 Strengths and Limitations

A major strength of this review is its multidimensional assessment of ASD diagnostic access, incorporating diagnostic completion, pathways, and timely diagnosis, alongside complementary frequentist and Bayesian analyses. However, several limitations should be considered. Only nine unique studies contributed quantitative data, limiting the precision of some subgroup and meta-regression estimates. Differences in study designs, populations, outcome definitions, and effect measures contributed to substantial heterogeneity. Multiple effect measures also mean that pooled estimates should be interpreted as standardized summary estimates rather than directly equivalent probabilities. Sparse or missing data resulted in the removal of some meta-regression coefficients, limiting interpretation of certain moderators. Finally, the evidence is predominantly from high-income settings, with limited evidence from Africa, restricting generalizability to low-resource contexts.

#### 3.8.10 Implications for Practice and Research

The findings suggest that improving ASD diagnostic access requires interventions across the entire diagnostic pathway, rather than focusing exclusively on screening. Health systems should strengthen developmental surveillance, referral coordination, provider training, family navigation, specialist capacity, and mechanisms for tracking children from referral to completed assessment. Navigation approaches may be particularly valuable for disadvantaged families facing multiple simultaneous barriers^12^.

Future research should prioritize longitudinal studies that measure specific stages of the diagnostic pathway separately and use standardized definitions of diagnostic delay and completion. More evidence is also needed from low- and middle-income countries, where shortages of specialists and diagnostic infrastructure may substantially alter the pathway. Recent evidence from Kenya, for example, demonstrates that children may encounter multiple healthcare contacts before diagnosis, with substantial delays between caregiver recognition and diagnostic confirmation ^28^.

### 3.9 Conclusion

Overall, this review demonstrates that access to ASD diagnostic services is substantial but highly variable, with approximately three-quarters of observed outcomes representing successful access. Families are most likely to successfully navigate diagnostic pathways and least likely to complete the diagnostic process, while timely diagnosis shows particularly large variation across studies. The findings indicate that family and socioeconomic circumstances, child developmental characteristics, provider recognition, and health-system organization interact to shape diagnostic access. Improving access therefore requires coordinated interventions that extend beyond screening to ensure that children identified as potentially autistic can progress efficiently from recognition and referral through to completed and timely diagnosis.

## Data Availability

All data produced in the present work are contained in the manuscript

---

**Supplementary Table S1.** Database search strategies.

| Database | Search strategy |
| --- | --- |
| **MEDLINE/PubMed** | ("Autism Spectrum Disorder"[Mesh] OR autism[tiab] OR ASD[tiab] OR autistic[tiab]) AND (diagnosis[tiab] OR "diagnostic services"[tiab] OR screening[tiab] OR "intervention services"[tiab] OR "treatment services"[tiab] OR "healthcare access"[tiab] OR "service availability"[tiab] OR "service utilization"[tiab] OR workforce[tiab] OR specialist*[tiab]) AND ("wait time*" [tiab] OR "waiting time*" [tiab] OR "diagnostic delay*" [tiab] OR "delay to diagnosis"[tiab] OR "age at diagnosis"[tiab] OR timeliness[tiab] OR "access delay*" [tiab]) |
| **Embase** | (autism OR autistic OR "autism spectrum disorder" OR ASD) AND (diagnosis OR "diagnostic services" OR screening OR "intervention services" OR "treatment services" OR "healthcare access" OR "service availability" OR "service utilization" OR workforce OR specialist*) AND ("wait time*" OR "waiting time*" OR "diagnostic delay*" OR "delay to diagnosis" OR "age at diagnosis" OR timeliness OR "access delay*") |
| **Scopus** | TITLE-ABS-KEY(autism OR autistic OR "autism spectrum disorder" OR ASD) AND TITLE-ABS-KEY(diagnosis OR "diagnostic services" OR screening OR "intervention services" OR "treatment services" OR "healthcare access" OR "service availability" OR "service utilization" OR workforce OR specialist*) AND TITLE-ABS-KEY("wait time*" OR "waiting time*" OR "diagnostic delay*" OR "delay to diagnosis" OR "age at diagnosis" OR timeliness OR "access delay*") |
| **Web of Science** | TS=(autism OR autistic OR "autism spectrum disorder" OR ASD) AND TS=(diagnosis OR "diagnostic services" OR screening OR "intervention services" OR "treatment services" OR "healthcare access" OR "service availability" OR "service utilization" OR workforce OR specialist*) AND TS=("wait time*" OR "waiting time*" OR "diagnostic delay*" OR "delay to diagnosis" OR "age at diagnosis" OR timeliness OR "access delay*") |
| **Grey literature** | Targeted searches using combinations of autism, "autism spectrum disorder", diagnosis, "diagnostic services", "healthcare access", "service availability", "diagnostic delay", "age at diagnosis", timeliness, workforce, and specialist. Reference lists of included studies and relevant reviews were also screened. |

